# End of Average. Understanding Overweight & Obesity: Rationale and Design

**DOI:** 10.64898/2026.06.05.26354975

**Authors:** Eva Vanbrabant, Jikke Hesen, Suzan Jordan, Codrin Mironiuc, Lotte HJM Lemmens, Yuliya Shapovalova, Gijs H. Goossens, Anne Roefs

## Abstract

**Background:** Obesity is globally recognized as a complex, multifactorial chronic disease, with biological, psychological, environmental and behavioural factors involved in both disease pathogenesis and maintenance. Although previous group-based studies demonstrated involvement of each of these factors, there is large inter-individual variability in the factors contributing to disease development as well as intervention outcomes, causing limited translatability to the individual level. This heterogeneity in treatment effectiveness might be due to differential causal and maintenance factors of obesity. To enable the transition from a one-size-fits-all approach to a more personalized approach for individuals with overweight or obesity, this study aims to investigate if and how the degree of weight loss and changes in daily life behaviour after a combined lifestyle intervention depend on individual baseline profiles comprising of person characteristics, biological, psychological, environmental and behavioural factors.

**Methods:** This study will include 600 individuals varying in BMI, 200 participants with a healthy BMI (18.5-24.9kg/m^2^), 200 with overweight (BMI 25.0-29.9kg/m^2^), and 200 with obesity (BMI ≥30.0kg/m^2^). For all participants, a comprehensive individual baseline profile is created, including person characteristics, biological, psychological, environmental and behavioural factors. A clustering method is applied to identify clusters of participants with similar characteristics. Next, we examine if and how these clusters are linked to bodyweight indicators measured at baseline, and how they relate to daily lifestyle behaviour, as measured by ecological momentary assessment (EMA) using a smartphone app and sensor technology (3-week measurements). Individuals with overweight or obesity will be randomized to the intensive lifestyle intervention or a lifestyle information condition, to determine if treatment response can be predicted based on cluster characteristics, how daily lifestyle behaviour changes after an intervention, and how changes in daily lifestyle behaviour relate to treatment response.

**Discussion:** The End of Average study aims to characterize a large set of individuals varying in body weight to predict intervention effectiveness measured as changes in body weight indicators and in daily lifestyle behaviours. If reliable predictors of treatment success can be identified, these can be applied in personalized lifestyle interventions to improve lifestyle behaviour, body weight management and overall health.

## 2. Introduction

Obesity is a global health issue, and is recognized as a complex, multifactorial, heterogeneous chronic disease that predisposes to a wide range of other non-communicable diseases and complications such as type 2 diabetes, liver disease, cardiovascular diseases, mental health problems, and certain cancers (1, 2, 3, 4,5, 6, 7). At the group level, obesity is known to be caused and maintained by biological, psychological, environmental and behavioural factors. On the individual level, there is large variability in the extent to which each of these variables contribute to the development of obesity and its complications (8). Currently, most research is done on a group level, comparing groups with and without overweight on these factors (7,8).

A group approach has been successful in identifying key risk, causal and maintenance factors for overweight and obesity. However, effect sizes are often small and variability within and across studies is large (9), suggesting that obesity is not only multifactorially determined but also that factors contributing to obesity differ across individuals (10, 11). For instance, it is well established that there is a link between depressive symptoms and obesity, but depressive symptoms were only found in 10-20% of individuals with obesity (12, 13, 14, 15). Thus, group-based findings do not translate well to an individual level.

Currently, the first line treatment for obesity is an intensive lifestyle intervention (according to international Obesity Guidelines, also see *Intervention description*} intervention description; 15, 16, 17, 18). A lifestyle intervention has been shown to be most effective if it contains dietary counselling, physical activity and cognitive behavioural therapy (15, 17). The effectiveness is about 8% weight loss after one year (15). However, there is large inter-individual variability in effectiveness (11, 15, 16, 19). It has for example been shown that BMI change after an intensive lifestyle intervention ranged from −25.4% to +5.0% (19). As a comparison, with the new generation of obesity medication up to 20% weight loss can be achieved (20, 21, 22), and on average 30% with bariatric surgery, one year post surgery (23).

However, weight regain often occurs after initial successful weight loss after a lifestyle intervention, only 20% of individuals remain weight stable for 1 year. Additionally, 50% of initial weight is regained after 2 years and more than 75% of the weight loss is regained within 5 years, but there is large heterogeneity in body weight trajectories (24, 25, 26, 27). Likewise, a Diabetes Prevention Programme showed that 70% of participants had regained the lost weight 7 years later (28, 29). Another study showed a regain of one-third of the lost weight after 1 year, and half of participants returned to their original weight after 5 years (30). Patients in an intensive lifestyle intervention program typically regain almost all weight after 4-5 years (15).

Other available options for obesity treatment also show high relapse rates. Weight regain is about 25-70%, 10 years after bariatric surgery (28, 31). Medication studies show that most participants had returned nearly to baseline weight within 10 years after treatment (28, 31). Similarly, recent studies on semaglutide and tirzepatide have shown quick weight regain after cessation of medication (20, 21, 22). Variation in intervention success and high relapse rates after treatment might be explained by the large individual variation in risk and maintenance factors of obesity, suggesting that a one-size fits-all treatment is not optimal and that personalized approach is needed (7, 16). Therefore, the End of Average study aims to create individual profiles based on person characteristics, biological, psychological, environmental and behavioural factors at baseline and to further investigate intervention effectiveness and changes in daily lifestyle.

### Objectives

The current End of Average study aims to characterize 600 individuals varying in BMI on different factors causing and maintaining obesity: person characteristics, biological, psychological, environmental and behavioural factors. Of these 600 individuals, 200 participants with a healthy BMI (18.5-24.9kg/m^2^), 200 participants with overweight (BMI 25.0-29.9kg/m^2^), and 200 participants with obesity (BMI ≥30.0kg/m^2^) are included (*1, 32*).

With detailed data on person characteristics, biological, psychological, environmental and behavioural factors, we will create individual baseline profiles that will be clustered based on similarity and examine whether these clusters relate to bodyweight indicators. We will investigate if these clusters of baseline profiles are linked to bodyweight indicators and daily lifestyle behaviour collected with ecological momentary assessment (EMA) at baseline. A group of healthy individuals are included to examine whether certain baseline profiles are more prevalent for people with overweight or obesity than for people with a healthy weight. This study includes a lifestyle information condition (INFO) and a group receiving an intensive lifestyle intervention (ILI) to assess variability in treatment success measured at post-treatment, at 6-month and at 12-month follow-up. Furthermore, if weight loss, maintenance or regain and changes in daily lifestyle can be predicted based on cluster characteristics established at baseline.

Note that the main aim of this study is *not* to show the average effectiveness of an intensive lifestyle intervention. Instead, we look beyond the average and aim to understand the degree of effectiveness on an individual level of an intensive lifestyle intervention as well as a lifestyle information condition.

### Trial design

#### Pre-registration

This study’s desired sample size, variables, hypotheses, and planned analyses were preregistered on AsPredicted (AsPredicted #156842) prior to any data being collected. EMA and Garmin codebook can be found on OSF (https://osf.io/8fvq4/). Furthermore, the study was registered on Research with Human Participants (CCMO)/Overview of medical research in the Netherlands under the reference NL-OMON53868 (https://www.onderzoekmetmensen.nl/nl/trial/53868).

#### Study participants

This study includes weight stable (+/− 3kg in 3 months) men and women aged 18-65 years old with a BMI above 18.5kg/m^2^ without any history of bariatric surgery, currently receiving any kind of weight loss treatment, current use of diabetes medication or currently receiving psychotherapy for any mental/physical condition.

#### Screening (phase 0)

Potential eligible study participants are asked to provide information about their body height, current/estimated bodyweight, age, sex, Dutch-speaking ability, highest completed education, pregnancy, working night shifts, smartphone possession and device type, possession of a digital weighing scale, previous and current body weight treatment, diabetes, other current medication, dietary allergies or intolerances, and other relevant health information upon online registration for the study. In case the above information meets the inclusion criteria, they will be contacted for a more detailed telephone screening. If all further inclusion criteria (see *Eligibility criteria*) are met, a date and time is scheduled to visit the research facility at Maastricht University (UM).

#### Measurement day (phase 1)

All participants (*n* = 600) complete phase 1 of the study. During the visit, several measurements will be performed, namely (1) body anthropometrics (BMI, WHR and body composition), (2) an oral glucose tolerance test (OGTT) to measure ghrelin, leptin, glucose, insulin, triglycerides and free fatty acids, (3) a 6-minute step test (6MST) to assess physical fitness, and (4) an extensive set of 14 validated questionnaires assessing for example diet motivation and mental health, and 2 additional questionnaires for personal characteristics and home address information to be able to characterize their daily living environment (see *Outcomes and Predictor Variables* + see EMA codebook OSF). These questionnaires are completed online at home within 3 days of the measurement day. Participants also invite three to five acquaintances (e.g., family, friends, colleagues) to complete the International Physical Activity Questionnaire (IPAQ) to get insight into the level of physical activity of people in the participant’s social environment. Additionally, they share their gender, age, body weight, height and relation to the participant (see EMA codebook OSF). All participants complete three weeks of Ecological Momentary Assessment (EMA) targeting lifestyle related variables (e.g., food intake, physical activity, emotions, etc) and wear a Garmin activity tracker (see *Outcomes and Predictor Variables*).

#### Intervention period (phase 2) and post-intervention measures (phase 3 and 4)

After the measurement day, participants with a BMI ≥ 25kg/m^2^ (n = 400) will be randomized to a 6-month intensive lifestyle intervention (ILI) or a lifestyle information condition (INFO). The INFO group receives a lifestyle book to independently work on their lifestyle for 6 months, the ILI group receives this book with an addition of guidance by a coach for six group and five individual sessions spread over 6 months (for more detail see *Intervention description*). Intervention adherence is registered for both groups. After completion of the 6-month intervention period, body weight and waist to hip circumferences are measured at home by the participants, and reported via an online questionnaire to the researchers, including a picture of them weighing themselves on the digital scales, with their feet and the digits indicating their weight visible. In addition, all participants complete a questionnaire on their use and evaluation of the lifestyle book (INFO and ILI) and are asked to provide an evaluation of their coach and coaching sessions (ILI only). The coach also evaluates the participant and coaching sessions (ILI only). Furthermore, after 6 and 12 months, body weight, waist and hip circumference and any changes in health, medication use, medical treatment, mental health treatment, and treatment for body weight are assessed with an online questionnaire. All participants with a BMI ≥ 25kg/m^2^ (n = 400) are also invited to participate in an intensive follow-up study that includes 6 months of EMA and wearing of the Garmin activity tracker. For further information on this study, see: AsPredicted.org #233961. See (3.) Methods and (4.) Interventions for a more detailed explanation of inclusion and measurement procedures.

## 3. Methods

### Study setting

The study is conducted at the university research facility, Maastricht University, Maastricht, the Netherlands (one-site study).

### Eligibility criteria

#### Phase 0: online screening

An online screening is completed as a first check of the eligibility criteria. Using an online (Qualtrics) questionnaire height, weight, age, highest level of (completed) education is asked and further inclusion criteria are checked. A participant is eligible if they are between 18 and 65 years old (on the day of inclusion), BMI > 18.5, speaks Dutch, resides in the Netherlands, does not have pregnancy wish within the coming 2 years and is not currently pregnant (or is not applicable), is in possession of a smartphone, is currently not receiving treatment for body weight, has not undergone gastric surgery, does not take diabetes medication, is not receiving treatment for any mental/physical condition, any medication for mental and physical disorder is stable (≥ 3 months) and does not have other health issues (possibly) interfering with study outcome. A participant might be eligible if days with night shifts occur less frequently than without night shifts, or if measurement day can be planned prior to 3 weeks without night shifts, if a digital weighing scale is not in their possession they will receive one for the duration of the study. Furthermore, treatment for body weight needs to be halted prior to study inclusion and need to be weight stable (treatment if medication needs to be halted 3 months prior to inclusion), other health issues are decided on a case to case basis. Subjects that are or might be eligible to participate in the study are asked for their contact details (email address and phone number), as well as first and last name, gender, preferred pronouns and availability for a screening phone call. See Table 1 below for a clear overview of eligibility criteria.

**Table 1:** Eligibility criteria.

| Criteria | Exceptions |
| --- | --- |
| ≥18 or ≤65 years old | NA |
| Speaks Dutch | NA |
| Resides in the Netherlands | NA |
| Weight stable for the past 3 months (+/- 3kg) | NA |
| Not pregnant or no short-term (2 year) pregnancy wish | Only applicable if BMI ≥ 25kg/m<sup>2</sup> |
| Not working regular night shifts | Exceptions if nights shift if days with night shifts occur less frequently than without night shifts, or if measurement day can be planned prior to 3 weeks without night shifts |
| In possession of a smartphone (& able to keep phone at reach) | NA |
| Has a digital weighing scale at home | Digital scale can be borrowed for duration of study period |
| Not currently receiving treatment for body weight | If treatment is halted 3 months before measurement day/start of study |
| No gastric surgery in the past | NA |
| No medication for diabetes type 1 or 2 | NA |
| Currently not under treatment for any mental/physical condition | NA |
| Other health issues | Decided on a case-to-case basis |

#### Phase 0: telephone screening

Eligible participants are contacted for a telephone screening. The researchers will then check all eligibility criteria and provide further study information. Potential study participants are given the chance to ask questions and to consider their study participation before they agree to participate.

A telephone screening includes registration of the date of the phone call and asking if the participant is able to come to the study site early in the morning. Demographics such as age, date of birth, and estimated height and weight to calculate BMI are asked. In- and exclusion criteria (see table 1 above) are checked again. If all eligibility criteria are met, a participant is assigned a screening number (s001-sxxx) and a visit is scheduled on the study site. Holidays of the participants are taken into account to ensure that the 3-week EMA period does not take place during their holidays as this will not be representative of daily lifestyle. Furthermore, it is considered if someone is a blood donor as blood donations should not be done in the 3 months prior to the visit. All participants are instructed to maintain weight stable until the measurement day (+/− 3kg) and participants with overweight and obesity are asked to weigh themselves the morning of the visit. All participants are provided with instructions for the measurement day, namely to be fasted for at least 12 hours, to not take any alcohol or drugs for 24 hours and to not engage in heavy exercise for 48 hours prior to the measurement day.

If a visit is planned more than three weeks ahead, a re-screening is planned one week prior to the visit to check if eligibility criteria are still met. Re-screening includes a check on weight stability (+/− 3kg), pregnancy status or wish, ongoing treatment for body weight, illness, other changes in health, current/past use of antibiotics, changes in medication, possession of a smartphone, and changes in night shift schedule. Table 2 gives an overview of the re-screening.

**Table 2:** Eligibility conditions checked in re-screening.

| **Condition to be met** | **Explanation?** |
| --- | --- |
| Weight stable for the past 3 months (+/- 3kg) | NA |
| Not pregnant or no short-term (2year) pregnancy wish | NA |
| Not currently receiving treatment for body weight | If yes, treatment needs to be halted before the measurement day, in case of medication 3 months prior to inclusion (medication stable) |
| Currently sick or sick in the past days | If yes, dates?<br>3 days recovered before visit* |
| Other changes in health since telephone screening | Decided on a case to case basis |
| Currently using antibiotics or in the past days/weeks | If yes, dates?<br>Antibiotics in the past 3 months = Excluded* |
| Medication stable for the past 3 months | If not, which medication was changed?<br>Decided on a case to case basis* |
| Still in possession of a smartphone | NA |
| Change in night shift working schedule | If yes; work night shifts more often than not OR working a planned nightshift the day before the visit* |

\*If not recovered, if antibiotics or medication are not stable, or if the night shift schedule is not optimal, a visit will be rescheduled or discussed with the participant.

#### Phase 1: Measurement day at UM

##### Who will take informed consent?

The informed consent form (ICF) is signed in duplicate on the measurement day on the study site by trained research staff and the participant.

##### Additional informed consent

Additional informed consent is given on the research day, see Table 3 below.

**Table 3:** Additional informed consent.

|  |  |  |
| --- | --- | --- |
| Permission to store data for other research. | Yes ☐ | No ☐ |
| Permission to store body materials for other research. Stored for 15 years. | Yes ☐ | No ☐ |
| Permission to be asked for a follow-up study. | Yes ☐ | No ☐ |
| Wants to be eligible for intensive follow-up of 6 months.* | Yes ☐ | No ☐ |

\*Not applicable for participants with a healthy BMI (18.5-24.9 kg/m<sup>2</sup>).

## 4. Interventions

### Explanation for the choice of comparators

For phase 2 (see fig. 1) participants are randomized to either an intensive lifestyle intervention (ILI) or a lifestyle information (INFO) condition for 6 months. Previous reviews have shown that an intensive lifestyle intervention is effective with an average weight loss of 9,3-10,9% (*15*) or up to 8kg (5-10%) weight loss after a 6-month intervention (*16*). These interventions included diet, physical activity and behavioural therapy and are comparable to the ILI of this study. Control conditions, like the INFO condition in the current study, only achieved 4,1-7,2% weight loss (*15*).

**Figure 1:**
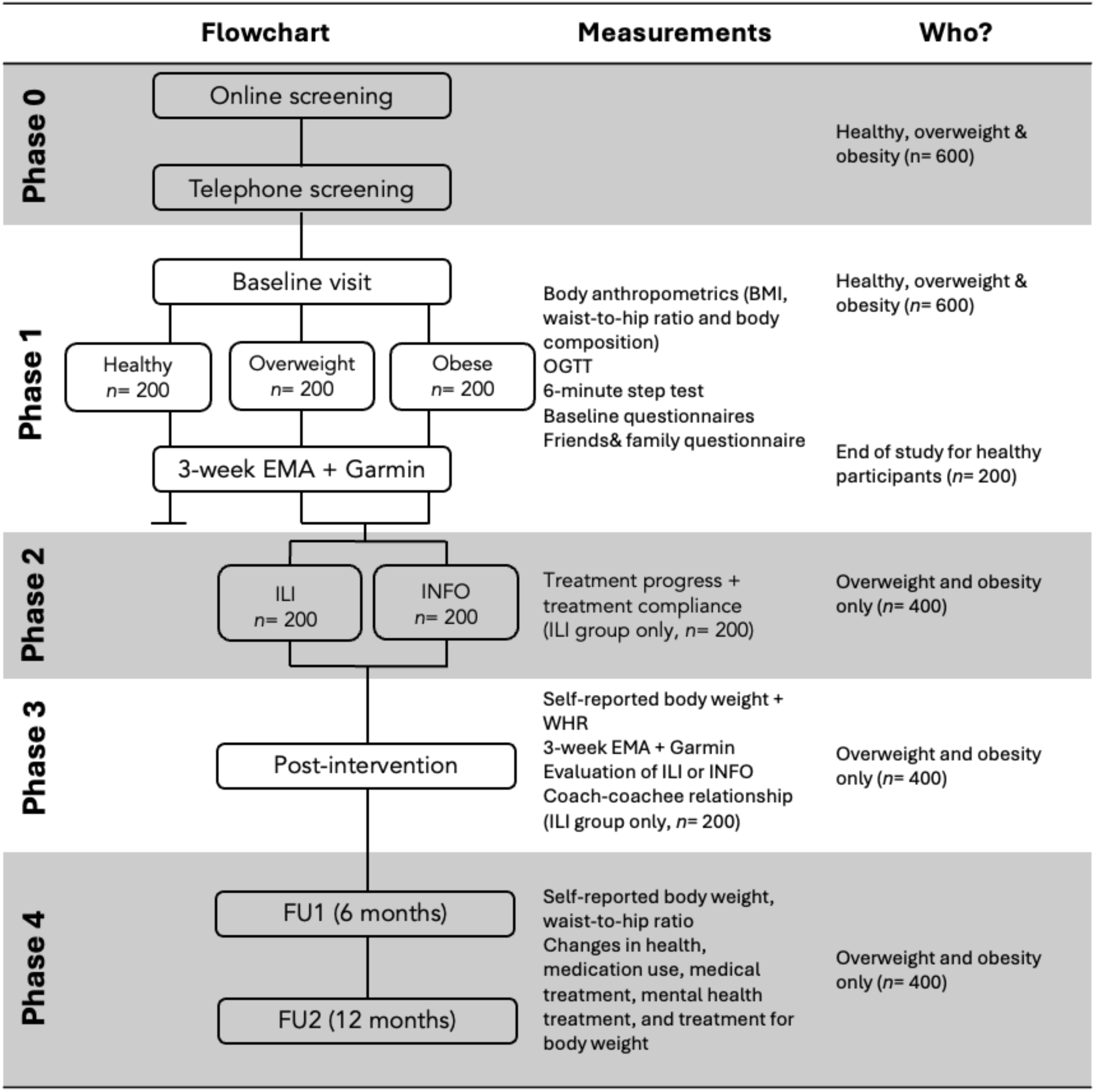
Flow chart overview of study including all different measurement phases. Part of this study is cross-sectional (phase 1) and part is a randomized controlled trial (phase 2, 3 and 4). BMI, body mass index; OGTT, oral glucose tolerance test; WHR, waist-to-hip ratio; EMA, ecological momentary assessment.

### Intervention description

#### Intensive lifestyle intervention (IL)

Participants allocated to the ILI group will participate in six group sessions and five individual coaching sessions, and are given three possibilities to ask questions via email communication, over 6 months provided online by a trained FIT.nl coach, and guided by the ‘SLANKER’’ handbook (33). All FIT.nl coaches are professional, certified coaches, dietitians and/or personal trainers. The intervention is based on cognitive behaviour (CBT) principles developed in our laboratory and are also in line with intensive lifestyle treatment guidelines (15, 16, 17, 33). The groups sessions are structured as follows:

Session 1: Introduction discussing motivation, expectations and goal setting
Session 2: Explaining CBT model
Session 3: Influence of emotions and behaviour
Session 4: Physical activity
Session 5: Environmental influences
Session 6: Maintaining (new) healthy lifestyle and relapse prevention

Individual sessions are tailored to the needs of the participant. The lifestyle coach keeps track of what is addressed in each individual session. Coaches record an evaluation of the overall well-being of the participant, homework completion, which problems, challenges or themes that were discussed (e.g., knowledge on healthy food, causes of relapse, dieting, sabotaging thoughts), and the interventions or strategies that were discussed (e.g., setting of realistic goals, planning food and exercise ahead of time, avoiding difficult/tricky situations). The coach also records any (planned) email contact or any other relevant information. Three contact moments with participants over the phone or via email are advised. For group sessions, a process measurement is completed after every group session including which specific sub-topics of a group session were addressed and if any other information is relevant for the research team.

#### Lifestyle information condition (INFO)

Participants allocated to the INFO group will receive the FIT.nl handbook on a healthy lifestyle called ‘SLANKER’ (translated to ‘LEANER’) to work with independently for 6 months. The ‘SLANKER’ handbook includes chapters on goal setting, behaviour change, nutrition and physical activity (33). Participants are advised to work with this handbook individually.

### Criteria for discontinuing/modifying allocated interventions

Criteria for discontinuing the allocated intervention are any adverse events, voluntary withdrawal of the participant, or any changes in medication or health status that can interfere with the study intervention or outcomes. Modifying the intervention is not possible (see *Intervention description*). Rare cases are discussed on a case to case basis, for example switching of coaches for certain sessions (ILI only).

### Strategies to improve adherence to interventions

Intervention adherence is recorded by the coaches and checked by the researchers, and, for the ILI group, make-up assignments are provided to participants if group sessions are missed. Make-up assignments include a video recording of the sessions and a homework assignment to ensure all participants complete the same (homework) tasks.

### Relevant concomitant care permitted or prohibited during the trial

Participants are excluded if any other treatment or obesity management medication will be initiated throughout the baseline, ILI or INFO period and follow-up measurements. During the follow-up period participants can undertake weight loss initiatives but are asked to report these in the 6 and 12 month post-intervention measurements if applicable.

### Provisions for post-trial care

All participants in the INFO group receive the option for a free intake with FIT.nl after finishing the 6-month INFO period and the 3-week post intervention measures. This study does not provide additional post-trial care.

### Outcomes and Predictor Variables

The main outcomes of the study, namely (self-measured corrected) body weight to calculate BMI and waist-to-hip ratio, will be measured at four time points: at baseline (phase 1), at the end of the intervention (phase 3), after 6 months and after 12 months of follow-up (phase 4). Furthermore, as a secondary outcome measure at baseline and after completion of the intervention, 3-week daily lifestyle measurements are performed with EMA and a Garmin activity tracker. These measurements are described in more detail below.

This study also includes biological, psychological, environmental, and behavioural measures that are not outcome variables, but that are predictor variables which are essential for testing our main research questions regarding the relationship and clustering between multifactorial comprehensive baseline profiles and body weight (change), as mentioned in section *Objectives*. The selection of these variables was based on the Accumulating Data to Optimally Predict Obesity Treatment (ADOPT) Core Measures Project implemented by The Obesity Society (8). This framework provides an integrated model including factors in the behavioural, biological, environmental and psychological domain that may influence obesity causes and treatment response (8, 35, 36, 37, 38). The ADOPT approach aims to improve efficacy of treatment by aiming to understand individual variability in treatment response by taking into account behavioural, biological, environmental and psychosocial influences, and using these as predictors of weight regain (7, 8, 15, 39).

#### Body Mass Index and Waist-to-Hip Ratio

Body weight and height are measured in duplicate without shoes and heavy clothing. Body weight is measured to the nearest 0.1kg using a weighing scale that is calibrated weekly. Waist and hip circumference are measured to the nearest 0.1 cm using a non-flexible measuring tape. All manual measurements (eg. height and waist and hip circumference) are measured three times, and an average is calculated. Measurement difference for height and WHR should not exceed 1cm. If measurement differences exceed 1cm, measurements are repeated. At baseline, body height, weight and waist-to-hip ratio are measured by the researchers as per the respective standard operating procedure (SOP).

After the intervention, participants self-report these measurements to the researchers following the same SOP. Participants are also asked to weigh themselves at home the morning of the measurement day to calculate the measurement difference between their (digital) home scale and the weighing scale on the test site. For all follow-up measurements of body weight, the measurement error is calculated with a percentage error. For home measurements that will take place at the end of the intervention and at follow-up, the participants are required to send a picture of them weighing themselves on the digital scales, with their feet and the digits indicating their weight visible.

#### Body Composition and Resting Energy Expenditure

Body composition is measured using the BodPod which uses the principle of Air Displacement Plethysmography. This allows for measurement of resting metabolic rate, percentage fat mass (%), percentage fat free mass (%), fat mass (kg) and fat free mass (kg) (40). Resting Energy Expenditure (REE) is also calculated with the Mifflin-St. Jeor equation (41, 42, 43). The Mifflin-St. Jeor equation gives a reliable estimate of RMR to within 10% of that measured out of the four available predictive equations (Mifflin-St. Jeor, Harris-Benedict, Owen, WHO/FAO/ONU (World Health Organization/Food and Agriculture Organization/United Nations University)) (42, 43).

#### Oral glucose tolerance test

A 2-hour, 5-point oral glucose tolerance test (OGTT) will be performed (*44*), with blood sampling taking place at 5 time points. Briefly, subjects ingest 200 ml of a ready-to-use 75 g glucose solution (Novolab) within 5 min, and blood samples are collected from the antecubital vein via an intravenous cannula under fasting conditions (*t* = 0 min) and after ingestion of the glucose drink (*t* = 30, 60, 90 and 120 min) for determination of fasting and postprandial plasma glucose, insulin, free fatty acids and ghrelin concentrations, leptin and triglyceride concentrations will measured in fasting state (*t* = 0).

#### Six-minute step test (Physical fitness test)

The 6-minute step test (6MST) is a highly accessible tool to assess cardiorespiratory fitness for a population varying in age and body weight (45, 46). Using a fitness step (height = 15cm) participants are instructed to step on and off the step as often and as fast as possible for 6 minutes. Before the test, resting heart rate is measured with a Garmin Vivosmart 4, and leg fatigue and dyspnea are measured using the scale for the rating of perceived exertion (Borg CR-10 scale; 0 - 10). During the test steps are counted and heart rate is recorded every minute using the same Garmin device, and standardized motivational sentences are given every minute. Leg fatigue, dyspnea and heart rate are assessed again 1-minute post-test using the same scale (45, 46).

#### Baseline questionnaires (see EMA codebook OSF)

All participants are instructed to complete an extensive online (Qualtrics) questionnaire which takes approximately 45-60 minutes to complete, within three days of the measurement day at UM. This questionnaire includes 14 validated surveys for psychological, behavioural and personal variables, and 2 additional questionnaires:

- Personal characteristics (age, highest completed level of education, employment status, monthly (household) income, sex, ethnicity, age of onset of overweight and/or obesity, parental BMI (father and mother: current and highest BMI; mother: highest BMI during pregnancy), menstrual cycle (if applicable), smoking status, alcohol and drug consumption, medication use, and possible other health issues.)
- Beck’s depression inventory II (BDI-II) (47, 48)
- Power of Food Scale (PFS) (49)
- Barratt Impulsiveness Scale-11 (BIS-11) (50)
- Short Self-Regulation Questionnaire (SSRQ) (51)
- Eating Disorder Examination Questionnaire 6.0 (EDE-Q) (52, 53)
- Dutch Eating Behaviour Questionnaire (54)
- Dichotomous Thinking in Eating Disorders Scale (DTEDS-11) (55)
- Body Image Concern Inventory (BICI) (56, 57)
- Rosenberg Self-Esteem Scale-11 (58, 59)
- Behavioural Regulation of Exercise Questionnaire (BREQ-2) & Regulation of Eating Behaviour Scale (REBS) (60, 61, 62)
- Health Status/QoL - Euroqol-5 (63)
- Home Address
- Social Support and Eating Habits + Social Support and Exercise Survey (64)
- Exercise Vital Sign Questionnaire (65)
- PrimeScreen (66)

At the end of the questionnaire, email addresses of three to five family members, close friends, or colleagues are asked to get an insight into the social environment of the participant. They were asked to share their relation to the participant, gender, age, body weight and height and complete the International Physical Activity Questionnaire (IPAQ) to get insight into the level of physical activity of people close to the participant (see EMA codebook OSF) (67).

#### Garmin Activity Tracker

After baseline measures at UM are completed, participants will complete three weeks of daily lifestyle measures using a Garmin Vivosmart 4/5 activity tracker to track sleep, physical activity and heart rate continuously (see EMA codebook OSF).

#### Ecological Momentary Assessments

Participants complete three-weeks of Ecological Momentary Assessment (EMA) (68) using the Avicenna Research application on their own smartphone. The app will trigger nine questionnaires per day. The ‘Sleep survey’ is triggered once a day at 07:00am, asking about sleep duration, sleep quality and nightly eating behaviour. Eight ‘Lifestyle surveys’ are semi-randomized every two hours. This survey contains questions about current mood states, where and with who the participant is at that moment, any eating moments since the last beep with the accompanying emotion, where, with whom, and how much someone ate, as well as physical activity duration, type, and intensity since the last beep. On the last day of this three-week study period, participants complete a short evaluation questionnaire to assess the perceived burden of the EMA study and whether (and how) the measurements affected their lifestyle behaviour (see EMA codebook OSF).

#### Measurements during the intervention period (see phase 2 figure 1, or figure 2)

Participants with overweight (*n* = 200) and obesity (*n* = 200) will be allocated to the ILI or INFO intervention groups (see *Intervention description* for more detailed explanation). For participants randomized to the ILI group, intervention progress is tracked by using treatment adherence forms that are completed by the coaches after every individual or group session. For the INFO group, adherence and book use is assessed by asking participants to fill in a short questionnaire at the end of the 6-month period. The same questions are also asked to the participants allocated to the ILI group, in addition to questions to evaluate the individual and group coaching sessions, and their lifestyle coach. For participants in the ILI condition, the coach also evaluates the participant, and the participant the coach, using a coach-coachee relationship questionnaire based on a short, revised form of the working alliance inventory (WAI-SR) (69) (see EMA codebook OSF).

**Figure 2:**
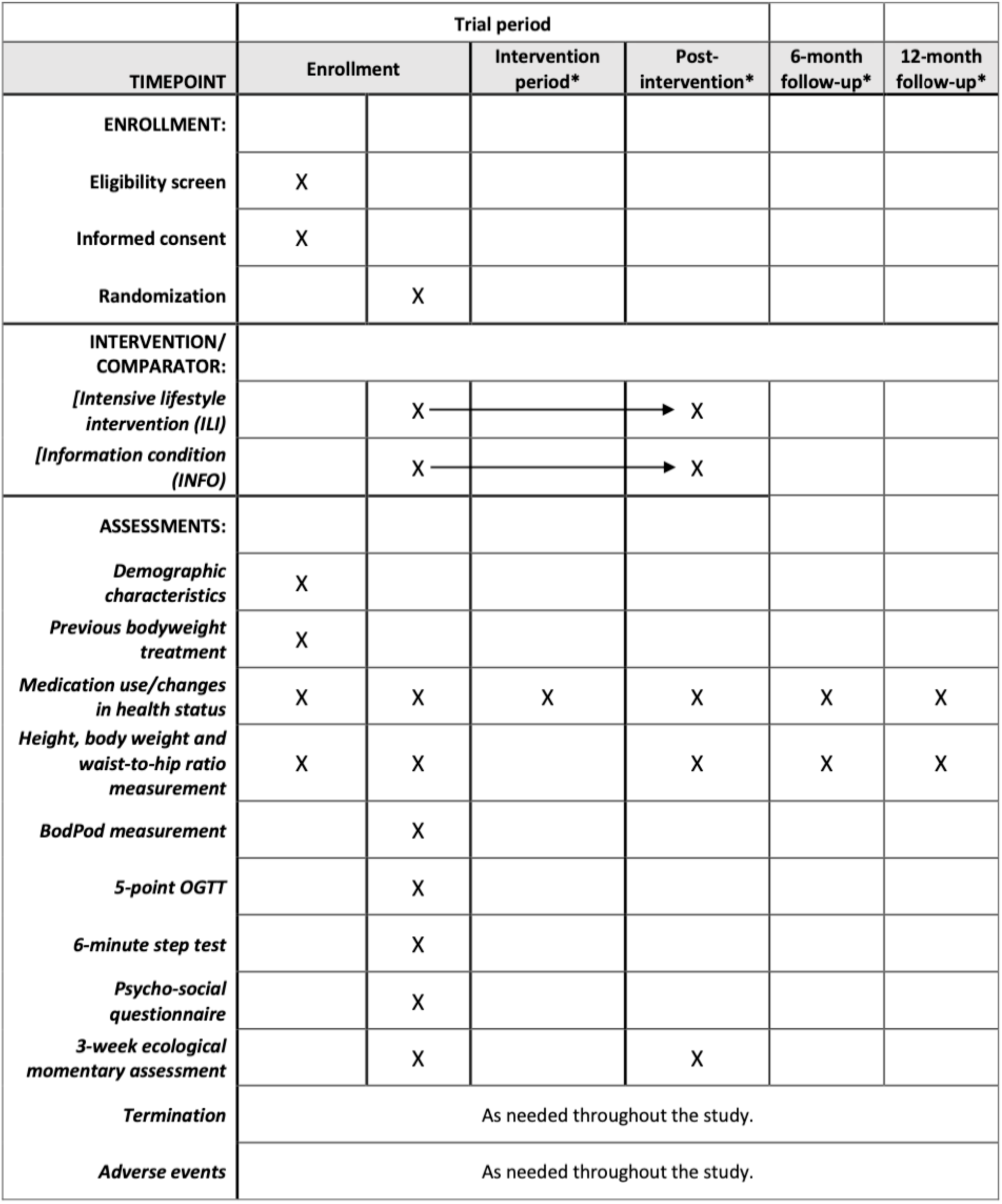
SPIRIT figure randomized controlled trial as a timeline of study participation. *\*Intervention period, post-intervention and follow-up measures only apply to participants with overweight or obesity*.

#### *Post-intervention measures* (see phase 3 figure 1, or figure 2)

After the six-month intervention, all participants (*n* = 400) self-measure their body weight and WHR. Participants in the ILI condition are asked to measure their bodyweight the day after the last coaching session. This can be a group or individual session, whichever comes last. Participants in the INFO condition are asked to measure their body weight 210 days after the baseline visit. This questionnaire includes questions specific to the intervention condition (as explained above), as well as questions about any medication, health changes or pregnancy (if applicable). Additionally, a repetition of 3-week EMA and Garmin-wearing is completed (see above for more detailed explanation).

### Follow-up measurements (see phase 4 figure 1, or figure 2)

After 6 months and 12 months of follow-up after the intervention has ended, self-reported body weight, waist and hip circumferences, as well as changes in health, medication, and pregnancy (if applicable), and any other relevant information such as (additional) weight loss attempts will be assessed.

### Participant timeline

**Figure 3:**
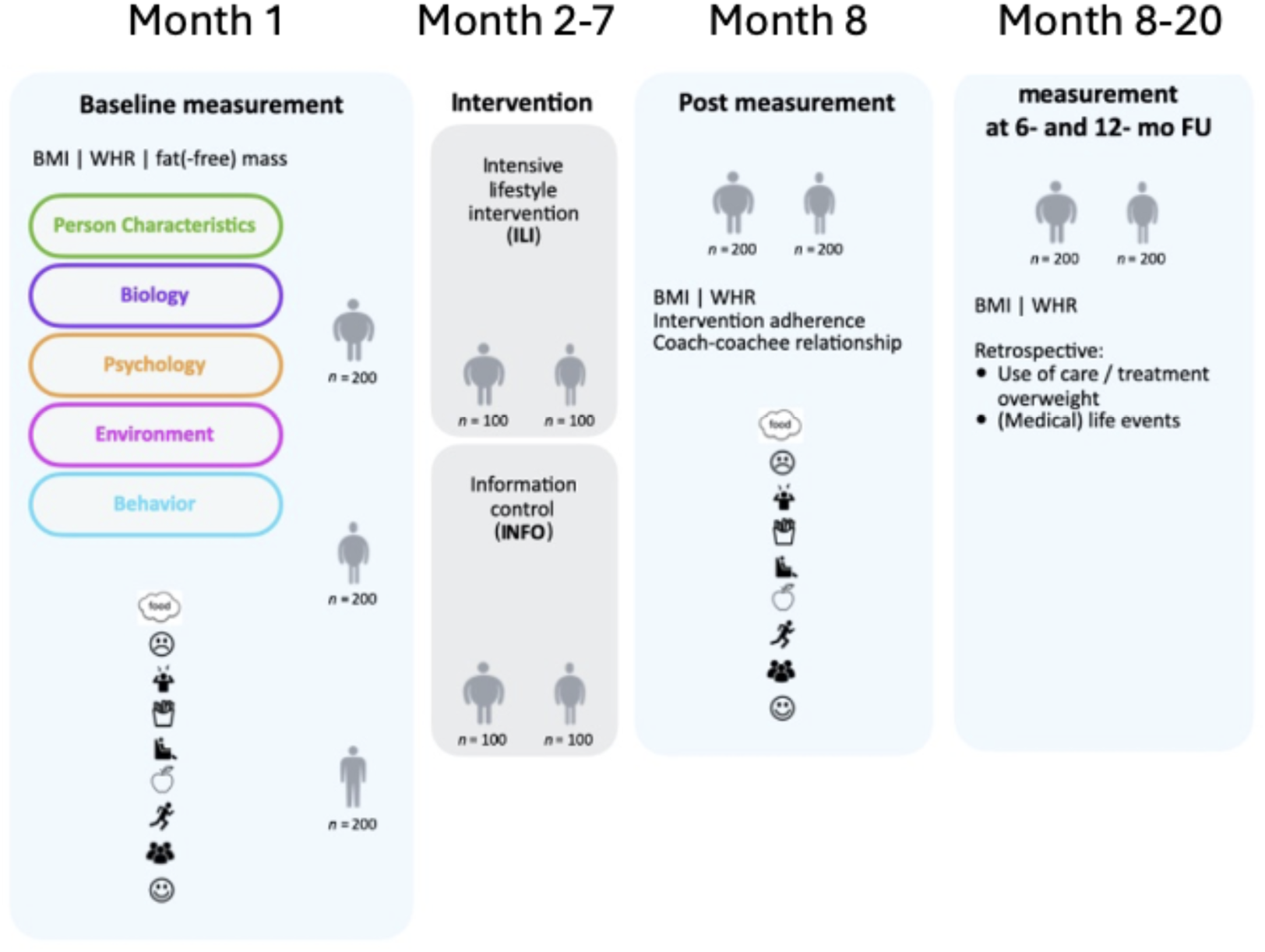
Timeline of study providing an overview of the measurements at baseline, during the intervention and at follow-up. BMI, Body Mass Index; WHR, waist-to-hip ratio; FU, Follow-up.

### Sample size

A sample of 600 participants is required for this study, of which 200 with a healthy weight (BMI 18.5-24.9kg/m^2^), 200 with overweight (BMI 25.0-29.9kg/m^2^) and 200 with obesity (BMI > 30.0kg/m^2^). A sample size of 600 participants is needed to give sufficient power for cluster analysis (70), also when allowing for 25% attrition. Note that attrition was 12% in a previous study of our lab with an intensive Ecological Momentary Assessment (EMA) component (71). During recruitment, we aim to balance person characteristics such as gender, education level, and/or age across bodyweight groups.

### Recruitment

Recruitment started in October 2023. Participants are recruited via our own social media channels (LinkedIn, Facebook, and Instagram), via general practitioners, flyer and poster distribution, via (local) newspapers/magazines and media outlets, the members network of FIT.nl, media appearances of members of the research team, and through (local) sport clubs.

## 5. Assignment of interventions: Allocation & randomization

### Sequence generation

Participants with overweight and obesity (*n* = 400) are randomized using Randomization.com. Four-hundred participants are randomized into 20 blocks of 10 ILI and 10 INFO participants per block. Randomization was done on 23/10/2023. The randomization plan can be reproduced on Randomization.org using seed *19527*, the number of participants per block (20) and the number of blocks (20). An extension was created on 12/08/2025 to randomize two more blocks of 20 of 10 ILI and 10 INFO (*n* = 40) to account for a possible 20 extra participants with overweight and 20 extra participants with obesity to compensate for possible drop-out replacement.

### Concealment mechanism

Randomization was finalized at the start of the study, and a hard copy is available on site. The allocated participant ID (EoA001 - EoA600) is filled out on the randomization list on the measurement day with a subsequent randomization number for treatment allocation. Participants are consecutively given a subject participation number as they visit (EoA001 - EoA600), and participants with overweight and obesity are allocated to ILI or INFO (overweight = randomization numbers 1-200, obesity = 201 to 400). Randomization list is digitalized on our data management server at regular intervals.

### Implementation

The allocation sequence was generated using Randomization.com as stated above. Participants are enrolled by trained researchers. Assignment of participants to the intervention is done by trained researchers.

## 6. Assignment of interventions: Blinding

- Who will be blinded & Procedure for unblinding if needed

Due to the nature of the intervention, blinding is not applicable.

## 7. Data collection and management

### Plans for assessment and collection of outcomes

All measurements are performed by the primary researchers or trained (student) researchers. Data quality is assured by repeating measures if necessary (as described in *Outcomes and Predictor Variables*). The respective SOPs of our laboratory are used. All included questionnaires are widely used and have sufficient to good reliability and validity (see *Outcomes and Predictor Variables* for Psychological questionnaires and EMA codebook OSF).

### Plans to promote participant retention and complete follow-up

#### Ecological Momentary Assessment

Compliance with the EMA protocol (i.e., the completion of surveys in the Avicenna Research app) is checked daily. Notification reminders are sent if compliance drops below 65% surveys completed on Monday through Thursday. On Fridays, a stricter cut off of 70% is used for compliance notifications because on weekends (Saturday and Sunday) compliance is not checked by researchers. Compliance reminders are standardized as an in-app notification and an additional email. If compliance is not increasing, a phone call is made to the participant to ensure the participant receives survey notifications and is able to keep up with survey load. Halfway through the 3-week period, a standardized email is also sent to the participants to notify them about the remainder of the study period. This procedure is the same for the baseline and post-intervention EMA study period. For data analysis, a minimum number of completed prompts is needed therefore compliance is checked daily.

#### During intervention period

All participants in the ILI and INFO group receive an encouraging postcard halfway the intervention period to motivate them. After completion of all measurements, a ‘thank you’ postcard is sent to participants to express our gratitude for their participation.

All emails for measurements are scheduled and sent using an in-house logistics research system (SOTO). This system also sends standardized reminders to participants and researchers if measurements are not completed. Researchers contact the participants via telephone or email if measurements are not completed despite SOTO-triggered reminders.

### Data management

For all data collection and storage, the study adheres to EU GDPR guidelines. Only one locked identification file links participant ID (EoAxxx) to screening number (sxxx). The screening number is linked to contact details in another locked file. These files are saved on a private data management server that only authorized researchers can access via a double authenticator method (also see *Confidentiality* below). All data during a test day is collected on paper case-report forms (CRF) and digitized to Castor EDC the same day. All paper CRFs are safely stored in a locked cabinet in the researcher’s office. Documents containing measurements collected by the researcher (e.g., CRFs, BodPod results, OGTT supervision form and pipetting scheme) only contain the study ID and are stored separately from documents containing personal or identifying information (e.g., screening forms, ICF, reimbursement form, Garmin contract and weighing scale contract (if applicable). All (non-private) data is digitalized on Castor, an Electronic Data Capture Software for Clinical Trials. See appendix for complete data management plan.

### Confidentiality

All participants are assigned a study ID according to the End of Average 001 to 600, abbreviated to EoA001 to EoA600. If needed, drop outs are replaced and study numbers > EoA600 are used. As stated above *Data management*) no personal identifying information is stored together with participant IDs.

### Plans for collection, laboratory evaluation and storage of biological specimens for genetic or molecular analysis in this trial/future use

Blood processing is done according to the respective SOP available in our laboratory. All samples are stored at −80℃. Samples are temporarily stored on site and stored at the biobank of the Maastricht University Medical Centre (MUMC) for long-term storage. Samples are stored for up to 15 years (see Table 5). On the ICF, permission is asked to use samples for other or future research (see *Additional informed consent* and Table 5).

## 8. Statistical methods

### Statistical methods for primary and secondary outcomes and predictor variables

#### 1. Individual profiles/baseline data & clustering

Different clustering algorithms are explored to cluster individuals with similar comprehensive profiles, implemented in R. For example, K-means clustering, polythetic agglomerative hierarchical clustering (PAHC), and Gaussian Mixed Models. If new methods develop over the course of this project, these methods will be explored as well. The aim is to identify a level of clustering that maximizes within-group similarity and minimizes among-group similarity.

Robustness and reproducibility of identified clusters will be tested, for example using Dunn’s Index. Identified clusters will be compared on different body weight indicators (BMI, WHR, Bodyfat percentage), and lifestyle at different timepoints (baseline, post-intervention and follow up). For comparison at baseline body weight, all participants (n = 600) are included in the cluster analysis. The cluster analysis is then repeated for the participants with a BMI > 25 (n = 400) to find associations between identified clusters and treatment outcome (BMI, WHR and lifestyle).

#### 2. Time-series data

For the time-series data, we consider using a vector autoregressive model (VAR) to estimate both contemporaneous (associations at 1 timepoint) and temporal (e.g., prediction from timepoint t-1 to t) daily lifestyle networks. For the temporal networks, each variable is regressed on a time-lagged version of that same variable and all the other variables in the network. The residuals of the VAR model will determine the contemporaneous models, after temporal associations have been controlled for. Cluster differences on network characteristics (i.e., node average and edge strength) will be tested using MANOVA. In addition, a clustering analysis is performed on the time-series data and will proceed similarly as with the clustering analyses of the comprehensive baseline profiles specified above.

#### 3. Prediction models

Furthermore, to test the prediction that a healthier lifestyle network is associated with a lower bodyweight, the levels of healthy and unhealthy behaviours (node averages) are extracted from the estimated networks, as well as the strengths of the edges connecting predictor variables (e.g., emotions) to these behaviours. Next, scores on network characteristics (node sizes and edge strengths) are correlated with body weight. After checking multicollinearity, all network characteristics that are significantly correlated with bodyweight (p < 0.10) are added to a regression model predicting bodyweight. With this regression model, we test which network characteristics explain most variance in bodyweight. This analysis is performed both at baseline and post-intervention. Pre-to-post intervention changes in the daily lifestyle networks and moderation by treatment-condition are tested with a mixed MANOVA.

#### 4. State of art approach

The field of precision science is moving fast. Throughout the duration of the project the research team will keep a close eye on analytical developments in the fields of precision medicine, clustering analyses and time-series analyses. When better methods become available, they will be explored. Statistical methods are described now as pre-registered (AsPredicted, #156842). The dataset that we collect is very rich. Additional questions may be answered with this dataset as well, though they are not currently part of the main research program. Examples include the validation of Garmin data against self-reported activity data, and food intake across the menstrual cycle.

### Interim analyses

Interim analyses are done for conference presentations on a subset of the available data at the time of conference, but these analyses are not our final or main analyses.

Methods for additional analyses (e.g. subgroup analyses)

Stated above (see *Statistical methods* - state of art approach). As new techniques become available, the most suitable (statistical) analysis methods will be used, available at that moment in time.

### Methods in analysis to handle protocol non-adherence and any statistical methods to handle missing data

Participants who fail to complete the online baseline questionnaire or 3-week EMA measurement period at baseline are replaced. These data of these participants have too many missing observations and can therefore not be included in the cluster analysis.

For outliers in the clustering analysis, we will follow the suggestions put forward by Nowak-Brzezińska and Gaibei (2022) (72). This involves detecting outliers using outlier detection algorithms with different percent parameters before clustering and checking the quality of clusters with and without outliers.

The time-series data of daily lifestyle networks will be checked for careless responding, incomplete surveys, and other missing observations. Cases of careless responding and missing data are not automatically excluded but examined on a case-to-case basis. Adherence to minimal requirements will be checked for data completeness. For the estimation of VAR models, a minimum of ±50% completed prompts is required.

Further replacement of participants will be decided on a case to case basis, for example based on medication of health changes throughout the intervention period, or if time permits.

### Plans to give access to the full protocol, participant level-data and statistical code

Full protocol is available, and participant level data is available upon reasonable request.

## 9. Oversight and monitoring

### Composition of the coordinating centre and trial steering committee

The daily research at the coordinating research centre consists of one principal investigator (PI), three co-PI’s, three PhD students and one researcher. Support from student interns and assistants is available on a regular basis. The daily research teams meet once weekly with the PI, PhD students and researcher. The complete team meets monthly, and more frequently if needed. Furthermore, the project sounding board, including members from academia, knowledge centres and healthcare is updated yearly. Furthermore, the FIT.nl coaching team plays a major role in this research project as they are responsible for all the coaching of the ILI group participants and have written and published the ‘SLANKER’ handbook used by both the ILI and INFO group. Communication with the FIT.nl team and coaches is done as needed (via email, phone or Slack communication portal). The daily research team is responsible for all data management.

### Composition of the data monitoring committee, its role and reporting structure

Monitoring of the study takes place on-site by a fellow researcher from the faculty department who is not involved in any way in the research project. Three monitoring visits are planned where a verification of source documents is done, the trial master files are checked, and results are discussed and a report is written. A standardized monitoring form is available and is filled in during monitoring visits. The first monitoring visit was completed on 20/02/2025, the second will take place early 2026 when all baseline data is collected and the last will take place around July 2027 when all data collection is completed.

### Adverse event reporting and harms

Adverse events (AEs) are noted down on the measurement day CRF, as well as any SAEs. SAEs are reported according to guidelines to toetsingonline.nl.

### Frequency and plans for auditing trial conduct

See above.

### Plans for communicating important protocol amendments to relevant parties

As stated above (see *Composition of the coordinating centre and trial steering committee*), the complete research team meets monthly and the project sounding board is updated yearly. The Medical Ethics Review Committee of UM-MUMC+ is updated yearly as requested, through a template process report a year after approval and a report of included participants to the financial/insurance department/service of Maastricht University at the end of the calendar year.

### Dissemination plans

Results are shared with participants if participants wish to be informed about overall results at the end of the complete study. Participants are updated on the general study progress with a bi-yearly newsletter. Furthermore, participants are informed about their personal blood results and receive reports of each 3-week EMA data collection period. Results are shared with the scientific community via publications in scientific journals and conference presentations. The public is informed via the project website (www.theendofaverage.nl) and media appearances of the members of the project team.

## 10. Discussion

To our knowledge, this will be the first study to generate individual profiles based on personal, biological, psychological, environmental and behavioural factors in a large number of individuals varying in BMI to assess the relationship with body weight indicators and daily life behaviour, and to predict the effectiveness of a combined lifestyle intervention regarding changes in body weight indicators and daily lifestyle behaviours. In addition, we will assess daily lifestyle behaviours using both EMA and an activity tracker before and after an intensive lifestyle intervention in individuals with overweight and obesity, allowing for the assessment of changes in daily lifestyle behaviours. Altogether, the End of Average study will enable investigating the determinants of lifestyle intervention outcomes at the individual level, paving the way for personalized lifestyle strategies to improve lifestyle behaviour, body weight management and overall health in individuals living with overweight and obesity.

## 11. Trial status

Recruitment, participant testing and intervention are ongoing. Recruitment started on 13th June 2023 by allowing participants to show interest in participation. On 11th October 2023 participants could register online for screening by filling a short questionnaire (see *Trial design*). (Telephone) screening started on 22nd November 2023. Participant inclusion started on 8th January 2024. Participant inclusion is planned to end in December 2025. Data-collection will be complete by August 2028.

## Supporting information

Codebook

## Data Availability

Full protocol is available, and participant level data is available upon reasonable request.

## 12. Abbreviations

ADOPT: Accumulating Data to Optimally Predict Obesity Treatment
AE: Adverse Event
BDI-II: Beck’s depression inventory II
BIS-11: Barratt Impulsiveness Scale-11
BICI: Body Image Concern Inventory
BMI: Body Mass Index
BREQ-2: Behavioural Regulation of Exercise Questionnaire
CRF: Case Report Form
DTEDS-11: Dichotomous Thinking in Eating Disorders Scale
EDE-Q: Eating Disorder Examination Questionnaire 6.0
EMA: Ecological Momentary Assessment
EoA: End of Average
FU: Follow-up
ICF: Informed Consent Form
ILI: Intensive lifestyle intervention
INFO: Information group
IPAQ: International Physical Activity Questionnaire
METC: Medisch-ethische Toetsingscommissie (Medical Ethical Review Committee)
MUMC+: Maastricht Universitair Medisch Centrum+ (Maastricht University Medical Centre+)
OGTT: Oral Glucose Tolerance Test
PFS: Power of Food Scale
UM: Universiteit Maastricht (Maastricht University)
REBS: Regulation of Eating Behaviour Scale
REE: Resting Energy Expenditure
SAE: Serious Adverse Event
SOP: Standard Operating Procedure
SSRQ: Short Self Regulation Questionnaire
WHR: Waist-to-Hip Ratio

## 13. Declarations

## Acknowledgements

Not applicable.

## Authors’ contributions

Funding for the study was obtained by AR. AR, GG, LL, JH, SJ, CM and EV contributed to the design and methodology of the study. AR, LL, JH, SJ, CM and EV are responsible for the design of the EMA protocol. Project administration is done by JH, SJ, CM and EV. JH, SJ, CM and EV are responsible for recruitment and data collection. Data analysis will be carried out by JH, CM and EV in collaboration with AR, GG, LL and YS. Supervision is done by AR, GG, LL and YS. EV wrote the original draft preparation in collaboration with AR. Review and editing was done by AR, GG, LL, JH, SJ, CM and YS. The final version was written by EV and AR. The final manuscript was approved by all authors.

## Availability of data and material

Full protocol is available, and participant level data is available upon reasonable request.

## Ethics approval and consent to participate

This study was approved by the METC on 21/08/2023 with protocol number NL817110.068.23 and METC number METC23-020. The study is conducted according to the principles of the Declaration of Helsinki (revised version, 2013, Fortaleza, Brazil), and all subjects provide written informed consent before the start of the study.

## References

1. WHO (2025). “Obesity and overweight.” from https://www.who.int/news-room/fact-sheets/detail/obesity-and-overweight.

2. Afshin, A, et al. (2017). “Health Effects of Overweight and Obesity in 195 Countries over 25 Years.” N Engl J Med 377(1): 13–27. 10.1056/NEJMoa1614362

3. Wise J. (2021). Covid-19: Highest death rates seen in countries with most overweight populations. BMJ (Clinical research ed.), 372, n623. 10.1136/bmj.n623

4. Lingvay, I., Cohen, R. V., Roux, C. W. L., & Sumithran, P. (2024). Obesity in adults. *Lancet (London*, England), 404(10456), 972–987. 10.1016/S0140-6736(24)01210-8

5. Frühbeck, G., Busetto, L., Dicker, D., Yumuk, V., Goossens, G. H., Hebebrand, J., Halford, J. G. C., Farpour-Lambert, N. J., Blaak, E. E., Woodward, E., & Toplak, H. (2019). The ABCD of Obesity: An EASO Position Statement on a Diagnostic Term with Clinical and Scientific Implications. Obesity facts, 12(2), 131–136. 10.1159/000497124

6. Avila, C., Holloway, A. C., Hahn, M. K., Morrison, K. M., Restivo, M., Anglin, R., & Taylor, V. H. (2015). An Overview of Links Between Obesity and Mental Health. Current obesity reports, 4(3), 303–310. 10.1007/s13679-015-0164-9

7. MacLean, P. S., Wing, R. R., Davidson, T., Epstein, L., Goodpaster, B., Hall, K. D., Levin, B. E., Perri, M. G., Rolls, B. J., Rosenbaum, M., Rothman, A. J., & Ryan, D. (2015). NIH working group report: Innovative research to improve maintenance of weight loss. Obesity (Silver Spring, Md.), 23(1), 7–15. 10.1002/oby.20967

8. MacLean, P. S., Rothman, A. J., Nicastro, H. L., Czajkowski, S. M., Agurs-Collins, T., Rice, E. L., Courcoulas, A. P., Ryan, D. H., Bessesen, D. H., & Loria, C. M. (2018). The Accumulating Data to Optimally Predict Obesity Treatment (ADOPT) Core Measures Project: Rationale and Approach. Obesity (Silver Spring, Md.), 26 Suppl 2(Suppl 2), S6–S15. 10.1002/oby.22154

9. Hamaker, E. L. (2012). Why researchers should think “within-person“: A paradigmatic rationale. In M. R. Mehl & T. S. Conner (Eds.), Handbook of research methods for studying daily life (pp. 43–61). The Guilford Press.

10. Piccirillo, M. L., & Rodebaugh, T. L. (2019). Foundations of idiographic methods in psychology and applications for psychotherapy. Clinical psychology review, 71, 90–100. 10.1016/j.cpr.2019.01.002

11. Bomberg, E. M., Ryder, J. R., Brundage, R. C., Straka, R. J., Fox, C. K., Gross, A. C., Oberle, M. M., Bramante, C. T., Sibley, S. D., & Kelly, A. S. (2019). Precision medicine in adult and pediatric obesity: a clinical perspective. Ther Adv Endocrinol Metab, 10, 2042018819863022. 10.1177/2042018819863022

12. Luppino, F. S., de Wit, L. M., Bouvy, P. F., Stijnen, T., Cuijpers, P., Penninx, B. W., & Zitman, F. G. (2010). Overweight, obesity, and depression: a systematic review and meta-analysis of longitudinal studies. Archives of general psychiatry, 67(3), 220–229. 10.1001/archgenpsychiatry.2010.

13. Ziauddeen, H., Farooqi, I. S., & Fletcher, P. C. (2012). Obesity and the brain: how convincing is the addiction model?. Nature reviews. Neuroscience, 13(4), 279–286. 10.1038/nrn3212

14. Werthmann, J., Jansen, A., & Roefs, A. (2015). Worry or craving? A selective review of evidence for food-related attention biases in obese individuals, eating-disorder patients, restrained eaters and healthy samples. The Proceedings of the Nutrition Society, 74(2), 99–114. 10.1017/S0029665114001451

15. Webb, V. L. and T. A. Wadden (2017). “Intensive Lifestyle Intervention for Obesity: Principles, Practices, and Results.” Gastroenterology 152(7): 1752–1764. 10.1053/j.gastro.2017.01.045

16. Jensen, M. D., Ryan, D. H., Apovian, C. M., Ard, J. D., Comuzzie, A. G., Donato, K. A., Hu, F. B., Hubbard, V. S., Jakicic, J. M., Kushner, R. F., Loria, C. M., Millen, B. E., Nonas, C. A., Pi-Sunyer, F. X., Stevens, J., Stevens, V. J., Wadden, T. A., Wolfe, B. M., Yanovski, S. Z., Jordan, H. S., … Obesity Society (2014). 2013 AHA/ACC/TOS guideline for the management of overweight and obesity in adults: a report of the American College of Cardiology/American Heart Association Task Force on Practice Guidelines and The Obesity Society. Circulation, 129(25 Suppl 2), S102–S138. 10.1161/01.cir.0000437739.71477.ee

17. Werrij, M. Q., Jansen, A., Mulkens, S., Elgersma, H. J., Ament, A. J., & Hospers, H. J. (2009). Adding cognitive therapy to dietetic treatment is associated with less relapse in obesity. Journal of psychosomatic research, 67(4), 315–324. 10.1016/j.jpsychores.2008.12.011

18. Busetto, L., Dicker, D., Frühbeck, G., Halford, J. C. G., Sbraccia, P., Yumuk, V., & Goossens, G. H. (2024). A new framework for the diagnosis, staging and management of obesity in adults. Nature medicine, 30(9), 2395–2399. 10.1038/s41591-024-03095-3

19. Ryder, J. R., Kaizer, A. M., Jenkins, T. M., Kelly, A. S., Inge, T. H., & Shaibi, G. Q. (2019). Heterogeneity in Response to Treatment of Adolescents with Severe Obesity: The Need for Precision Obesity Medicine. Obesity (Silver Spring, Md.), 27(2), 288–294. 10.1002/oby.22369

20. Salvador, R., Moutinho, C. G., Sousa, C., Vinha, A. F., Carvalho, M., & Matos, C. (2025). Semaglutide as a GLP-1 Agonist: A Breakthrough in Obesity Treatment. *Pharmaceuticals (Basel*, Switzerland), 18(3), 399. 10.3390/ph18030399

21. Wilding, J. P. H., Batterham, R. L., Davies, M., Van Gaal, L. F., Kandler, K., Konakli, K., Lingvay, I., McGowan, B. M., Oral, T. K., Rosenstock, J., Wadden, T. A., Wharton, S., Yokote, K., Kushner, R. F., & STEP 1 Study Group (2022). Weight regain and cardiometabolic effects after withdrawal of semaglutide: The STEP 1 trial extension. Diabetes, obesity & metabolism, 24(8), 1553–1564. 10.1111/dom.14725

22. Horn, D. B., Linetzky, B., Davies, M. J., Laffin, L. J., Wang, H., Murphy, M. A., Zimner-Rapuch, S., Lau, E., Arad, A. D., & Lee, C. J. (2025). Cardiometabolic Parameter Change by Weight Regain on Tirzepatide Withdrawal in Adults With Obesity: A Post Hoc Analysis of the SURMOUNT-4 Trial. JAMA Intern Med. 10.1001/jamainternmed.2025.6112

23. Tettero, O. M., Monpellier, V. M., Janssen, I. M. C., Steenhuis, I. H. M., & van Stralen, M. M. (2022). Early Postoperative Weight Loss Predicts Weight Loss up to 5 Years After Roux-En-Y Gastric Bypass, Banded Roux-En-Y Gastric Bypass, and Sleeve Gastrectomy. Obesity surgery, 32(9), 2891–2902. 10.1007/s11695-022-06166-x

24. Anderson, J. W., Konz, E. C., Frederich, R. C., & Wood, C. L. (2001). Long-term weight-loss maintenance: a meta-analysis of US studies. The American journal of clinical nutrition, 74(5), 579–584. 10.1093/ajcn/74.5.579

25. Wing, R. R., & Phelan, S. (2005). Long-term weight loss maintenance. The American journal of clinical nutrition, 82(1 Suppl), 222S–225S. 10.1093/ajcn/82.1.222S

26. McGuire, M. T., Wing, R. R., & Hill, J. O. (1999). The prevalence of weight loss maintenance among American adults. International journal of obesity and related metabolic disorders : journal of the International Association for the Study of Obesity, 23(12), 1314–1319. 10.1038/sj.ijo.0801075

27. van Baak, M. A., & Mariman, E. C. M. (2025). Physiology of Weight Regain after Weight Loss: Latest Insights. Current obesity reports, 14(1), 28. 10.1007/s13679-025-00619-x

28. Wadden, T. A., Webb, V. L., Moran, C. H., & Bailer, B. A. (2012). Lifestyle modification for obesity: new developments in diet, physical activity, and behavior therapy. Circulation, 125(9), 1157–1170. 10.1161/CIRCULATIONAHA.111.039453

29. Bray, G. A., Kim, K. K., Wilding, J. P. H., & World Obesity Federation (2017). Obesity: a chronic relapsing progressive disease process. A position statement of the World Obesity Federation. Obesity reviews : an official journal of the International Association for the Study of Obesity, 18(7), 715–723. 10.1111/obr.12551

30. Butryn, M. L., Webb, V., & Wadden, T. A. (2011). Behavioral treatment of obesity. The Psychiatric clinics of North America, 34(4), 841–859. 10.1016/j.psc.2011.08.006

31. Bosworth, T. (2017). Weight recidivism after bariatric surgery: What constitutes failure?. MDedge. https://www.mdedge.com/diabeteshub/article/150969/obesity/weight-recidivism-after-bariatric-surgery-what-constitutes

32. Bowman-Busato, J., Schreurs, L., Halford, J. C. G., Yumuk, V., O’Malley, G., Woodward, E., De Cock, D., & Baker, J. L. (2025). Providing a common language for obesity: the European Association for the Study of Obesity obesity taxonomy. International journal of obesity (2005), 49(2), 182–191. 10.1038/s41366-024-01565-9

33. Van der Mark, J., Louwes, L., Smit, N., & Huizinga, E. (2023). SLANKER (3rd ed.). FIT.nl.

34. Lemmens, L. H. J. M., & Roelofs, J. (2018). Cognitieve gedragstherapie. In E. Simon, E. de Hullu, G. Smeets, & H. T. van der Molen (editors), Klinische Psychologie: Diagnostiek en behandeling (3 redactie, blz. 135-164). Noordhoff Uitgevers.

35. Rosenbaum, M., Agurs-Collins, T., Bray, M. S., Hall, K. D., Hopkins, M., Laughlin, M., MacLean, P. S., Maruvada, P., Savage, C. R., Small, D. M., & Stoeckel, L. (2018). Accumulating Data to Optimally Predict Obesity Treatment (ADOPT): Recommendations from the Biological Domain. Obesity (Silver Spring, Md.), 26 Suppl 2(Suppl 2), S25–S34. 10.1002/oby.22156

36. Sutin, A. R., Boutelle, K., Czajkowski, S. M., Epel, E. S., Green, P. A., Hunter, C. M., Rice, E. L., Williams, D. M., Young-Hyman, D., & Rothman, A. J. (2018). Accumulating Data to Optimally Predict Obesity Treatment (ADOPT) Core Measures: Psychosocial Domain. Obesity (Silver Spring, Md.), 26 Suppl 2(Suppl 2), S45–S54. 10.1002/oby.22160

37. Saelens, B. E., Arteaga, S. S., Berrigan, D., Ballard, R. M., Gorin, A. A., Powell-Wiley, T. M., Pratt, C., Reedy, J., & Zenk, S. N. (2018). Accumulating Data to Optimally Predict Obesity Treatment (ADOPT) Core Measures: Environmental Domain. Obesity (Silver Spring, Md.), 26 Suppl 2(Suppl 2), S35–S44. 10.1002/oby.22159

38. Lytle, L. A., Nicastro, H. L., Roberts, S. B., Evans, M., Jakicic, J. M., Laposky, A. D., & Loria, C. M. (2018). Accumulating Data to Optimally Predict Obesity Treatment (ADOPT) Core Measures: Behavioral Domain. Obesity (Silver Spring, Md.), 26 Suppl 2(Suppl 2), S16–S24. 10.1002/oby.22157

39. Luig, T., Anderson, R., Sharma, A. M., & Campbell-Scherer, D. L. (2018). Personalizing obesity assessment and care planning in primary care: patient experience and outcomes in everyday life and health. Clinical obesity, 8(6), 411–423. 10.1111/cob.12283

40. Lindsey, B. W., Shookster, D. E., Martin, J. R., & Cortes, N. N. (2021). Predictive Accuracy of the Nelson Equation via BodPod Compared to Commonly Used Equations to Estimate Resting Metabolic Rate in Adults. International journal of exercise science, 14(2), 1166–1177. 10.70252/TFJB1938

41. Mifflin, M. D., St Jeor, S. T., Hill, L. A., Scott, B. J., Daugherty, S. A., & Koh, Y. O. (1990). A new predictive equation for resting energy expenditure in healthy individuals. The American journal of clinical nutrition, 51(2), 241–247. 10.1093/ajcn/51.2.241

42. Frankenfield, D., Roth-Yousey, L., & Compher, C. (2005). Comparison of predictive equations for resting metabolic rate in healthy nonobese and obese adults: a systematic review. Journal of the American Dietetic Association, 105(5), 775–789. 10.1016/j.jada.2005.02.005

43. Cancello, R., Soranna, D., Brunani, A., Scacchi, M., Tagliaferri, A., Mai, S., Marzullo, P., Zambon, A., & Invitti, C. (2018). Analysis of Predictive Equations for Estimating Resting Energy Expenditure in a Large Cohort of Morbidly Obese Patients. Frontiers in endocrinology, 9, 367. 10.3389/fendo.2018.00367

44. Trouwborst, I., Gijbels, A., Jardon, K. M., Siebelink, E., Hul, G. B., Wanders, L., Erdos, B., Péter, S., Singh-Povel, C. M., de Vogel-van den Bosch, J., Adriaens, M. E., Arts, I. C. W., Thijssen, D. H. J., Feskens, E. J. M., Goossens, G. H., Afman, L. A., & Blaak, E. E. (2023). Cardiometabolic health improvements upon dietary intervention are driven by tissue-specific insulin resistance phenotype: A precision nutrition trial. Cell metabolism, 35(1), 71–83.e5. 10.1016/j.cmet.2022.12.002

45. Carvalho, L. P., Di Thommazo-Luporini, L., Aubertin-Leheudre, M., Bonjorno Junior, J. C., de Oliveira, C. R., Luporini, R. L., Mendes, R. G., Zangrando, K. T., Trimer, R., Arena, R., & Borghi-Silva, A. (2015). Prediction of Cardiorespiratory Fitness by the Six-Minute Step Test and Its Association with Muscle Strength and Power in Sedentary Obese and Lean Young Women: A Cross-Sectional Study. PloS one, 10(12), e0145960. 10.1371/journal.pone.0145960

46. Di Thommazo-Luporini, L., Pinheiro Carvalho, L., Luporini, R., Trimer, R., Falasco Pantoni, C. B., Catai, A. M., Arena, R., & Borghi-Silva, A. (2015). The six-minute step test as a predictor of cardiorespiratory fitness in obese women. European journal of physical and rehabilitation medicine, 51(6), 793–802.

47. Beck, A. T., Steer, R. A., Ball, R., & Ranieri, W. F. (1996). Comparison of Beck Depression Inventories-IA and-II in psychiatric outpatients. Journal of personality assessment, 67(3), 588–597. 10.1207/s15327752jpa6703_13

48. van der Does, A. J. W. (2002). BDI-II-NL. Handleiding. De Nederlandse Versie van de Beck Depression Inventory (2nd ed.). Lisse: Harcourt Test Publishers.

49. Lowe, M. R., Butryn, M. L., Didie, E. R., Annunziato, R. A., Thomas, J. G., Crerand, C. E., […], & Halford, J. (2009). The Power of Food Scale. A new measure of the psychological influence of the food environment. Appetite, 53, 114–118. 10.1016/j.appet.2009.05.016

50. Stanford, M. S., Mathias, C. W., Dougherty, D. M., Lake, S. L., Anderson, N. E., & Patton, J. H. (2009). Fifty years of the Barratt Impulsiveness Scale: An update and review. Personality and Individual Differences, 47, 385–395. 10.1016/j.paid.2009.04.008

51. Carey, K. B., Neal, D. J., & Collins, S. E. (2004). A psychometric analysis of the self-regulation questionnaire. Addictive behaviors, 29, 253–260. 10.1016/j.addbeh.2003.08.001

52. Berg, K. C., Peterson, C. B., Frazier, P., & Crow, S. J. (2012). Psychometric evaluation of the eating disorder examination and eating disorder examination-questionnaire: a systematic review of the literature. The International journal of eating disorders, 45, 428–438. 10.1002/eat.20931

53. Fairburn, CG & Beglin, SJ (2008). Eating Disorder Examination Questionnaire. In: Fairburn, C (2008). Cognitive Behaviour Therapy and Eating Disorders. Guilford Press. (Unpublished translation by Anita Jansen, Maastricht).

54. Van Strien, T., Frijters, J.E.R., Bergers, G.P.A., & Defares, P.B. (1986). The Dutch Eating Behavior Questionnaire (DEBQ) for assessment of restrained, emotional, and external eating behavior. International Journal of Eating Disorders, 5(2), 295–315. 10.1016/j.appet.2012.08.029

55. Byrne, S. M., Allen, K. L., Dove, E. R., Watt, F. J., & Nathan, P. R. (2008). *Dichotomous Thinking in Eating Disorders Scale-11 (DTEDS-11)* [Database record]. APA PsycTests. 10.1037/t48296-000

56. Littleton, H. (2005). *Body Image Concern Inventory (BICI)* [Database record]. APA PsycTests. 10.1037/t00353-000

57. van Rood, YR. & de Beurs, E. (2005). Nederlandse vertaling van de Body Image Concern Inventory. (H.L. Littleton,, 2005) Vertaald met toestemming van de auteur (H.L. Littleton,, 2005).

58. Rosenberg, M. (1979). Conceiving the Self. New York: Basic Books

59. Franck, E., De Raedt, R., Barbez, C., & Rosseel, Y. (2008). Psychometric properties of the Dutch Rosenberg Self-Esteem Scale. Psychologica Belgica, 48(1), 25–35. 10.5334/pb-48-1-25

60. Markland, D., & Tobin, V. (2004). A modification to the behavioural regulation in exercise questionnaire to include an assessment of amotivation. Journal of Sport and Exercise Psychology, 26(2), 191–196. 10.1123/jsep.26.2.191

61. Pelletier, L. G., Dion, S. C., Slovinec-D’Angelo, M., & Reid, R. (2004). Why do you regulate what you eat? Relationships between forms of regulation, eating behaviors, sustained dietary behavior change, and psychological adjustment. Motivation and emotion, 28(3), 245–277. 10.1023/B:MOEM.0000040154.40922.14

62. Helmink, J.H.M., Van Boekel, L.C., van der Sluis, M.E. & Kremers, S.P.J. (2011). Lange termijn evaluatie onder deelnemers aan de BeweegKuur: Rapportage van de resultaten van een follow-up meting bij deelnemers. Universiteit Maastricht

63. EuroQol Group (1990). EuroQol--a new facility for the measurement of health-related quality of life. Health policy (Amsterdam, Netherlands), 16(3), 199–208. 10.1016/0168-8510(90)90421-9

64. Sallis, J. F., Grossman, R. M., Pinski, R. B., Patterson, T. L., & Nader, P. R. (1987). The development of scales to measure social support for diet and exercise behaviors. Preventive medicine, 16(6), 825–836. 10.1016/0091-7435(87)90022-3

65. Coleman, K. J., Ngor, E., Reynolds, K., Quinn, V. P., Koebnick, C., Young, D. R., Sternfeld, B., & Sallis, R. E. (2012). Initial validation of an exercise “vital sign” in electronic medical records. Medicine and science in sports and exercise, 44(11), 2071–2076. 10.1249/MSS.0b013e3182630ec1

66. Rifas-Shiman, S. L., Willett, W. C., Lobb, R., Kotch, J., Dart, C., & Gillman, M. W. (2001). PrimeScreen, a brief dietary screening tool: reproducibility and comparability with both a longer food frequency questionnaire and biomarkers. Public health nutrition, 4(2), 249–254. 10.1079/phn200061

67. Craig, C. L., Marshall, A. L., Sjöström, M., Bauman, A. E., Booth, M. L., Ainsworth, B. E., Pratt, M., Ekelund, U., Yngve, A., Sallis, J. F., & Oja, P. (2003). International physical activity questionnaire: 12-country reliability and validity. Medicine and science in sports and exercise, 35(8), 1381–1395. 10.1249/01.MSS.0000078924.61453.FB

68. Shiffman, S., Stone, A. A., & Hufford, M. R. (2008). Ecological momentary assessment. Annual review of clinical psychology, 4, 1–32. 10.1146/annurev.clinpsy.3.022806.091415

69. Hatcher, R. L., & Gillaspy, J. A. (2006). Development and validation of a revised short version of the Working Alliance Inventory. Psychotherapy research, 16(1), 12–25. 10.1080/10503300500352500

70. Dalmaijer, E. S., Nord, C. L., & Astle, D. E. (2022). Statistical power for cluster analysis. BMC bioinformatics, 23(1), 205. 10.1186/s12859-022-04675-1

71. Boh, B., Jansen, A., Clijsters, I., Nederkoorn, C., Lemmens, L. H. J. M., Spanakis, G., & Roefs, A. (2016). Indulgent thinking? Ecological momentary assessment of overweight and healthy-weight participants’ cognitions and emotions. Behaviour research and therapy, 87, 196–206. 10.1016/j.brat.2016.10.001

72. Nowak-Brzezińska, A., & Gaibei, I. (2022). How the Outliers Influence the Quality of Clustering?. Entropy (Basel, Switzerland), 24(7), 917. 10.3390/e24070917

