## Supplementary material for "End of Average. Understanding Overweight & Obesity: Rationale and Design": Codebook

### End of Average – Codebook

### General information

This codebook includes all questionnaires and translations used in the End of Average: Understanding Overweight & Obesity project. For validated psychological questionnaires we do not reproduce the full list of items per scale but refer to the original scale, where possible we also provide a reference of the Dutch translation. If questionnaires were created or adapted for the purpose of this study, it is noted. For transparency and validation purposes, the full list of scale items can be shared upon reasonable request (see up-to-date contact information in the OSF description).

### Baseline survey

#### Personal characteristics

| Variable | Item | Response format |
|---|---|---|
| Age | [EN] What is your date of birth?<br>[NL] Wat is uw geboortedatum? | [EN] day/month/year<br>[NL] dag/maand/jaar |
| Marital status | [EN] Wat is your marital/civil status?<br>[NL] Wat is uw burgerlijke staat? | [EN] Married, unmarried, registered partnership, widowed, divorced<br>[NL] Gehuwd, ongehuwd, geregistreerd partnerschap, verweduwd, gescheiden |
| Highest level of education | [EN] What is your highest completed level of education?<br>[NL] Wat is het hoogste opleidingsniveau dat u hebt voltooid of de hoogste graad die u hebt behaald? | [EN] None, Elementary school, Secondary education (VMBO + HAVO), Secondary prepatory university education (VWO), Higher vocational education (MBO), Higher professional education (HBO), University education including PhD<br><br>[NL] Geen, basisonderwijs, VMBO, HAVO, VWO, MBO, HBO, universitaire opleiding inclusief PhD |
| Current living situation | [EN] What is your current living situation?<br>[NL] Wat is uw huidige leef-woonsituatie? | [EN] Alone, with partner, with your (own) family, student house/residential community, living at home (with parents), other namely {open text}<br>[NL] Alleenwonend, samenwonend met partner, samenwonend met (uw eigen) gezin, studentenhuis/ woongemeenschap, thuiswonend (bij ouders), anders, namelijk {open tekst} |
| Employment status | [EN] What is your current employment status? (multiple options possible)<br>[NL] Wat is uw huidige werksituatie? (meerdere opties mogelijk) | [EN] Full-time employment, part-time employment, on-call worker, unemployed, looking for a job, self-employed/entrepreneur, student, retired<br>[NL] Fulltime dienstverband, parttime dienstverband, oproepkracht, werkloos, werkzoekend, zelfstandig/ondernemer, student, gepensioneerd |
| Work sector | [EN] Which sector do you work in?<br>[NL] In welke sector werkt u?<br>*displayed if full-time, part-time, on-call or self-employed/entrepreneur is selected as employment status. | [EN] Healthcare and welfare; trade and services; ICT; justice, security and public administration; media and communication; education, culture, and science; technology, manufacturing and construction; |

| | | |
|---|---|---|
| | | tourism, recreation and hospitality; transport and logistics<br>[NL] Gezondheidszorg en welzijn; handel en dienstverlening; ICT; justitie, veiligheid en openbaar bestuur; media en communicatie; onderwijs, cultuur, en wetenschap; techniek, productie en bouw; toerisme, recreatie en horeca; transport en logistiek |
| Night shift work | [EN] Do you also work night shifts? How often do you work night shifts?<br>[NL] Werkt u ook nachtdiensten?; Hoe vaak werkt u nachtdiensten?<br>*displayed if full-time, part-time, on-call or self-employed/entrepreneur is selected as employment status. | [EN] Yes; No; Sporadically, once a month, weekly; 2-3 times a week; 4-5 times a week; more than 5 times a week<br>[NL] Ja; Nee; Sporadisch; een keer per maand; wekelijks; 2-3 keer per week; 4-5 keer per week; meer dan 5 keer per week |
| Monthly (household) income | [EN] What is the average gross monthly income of your household?<br>[NL] Wat is het gemiddelde bruto maandinkomen van uw huishouden? | [EN] < € 500; € 500 – € 1.000; € 1.000 – € 2.000; 2.000 – € 3.000; 3.000 – € 4.000; 4.000 – € 5.000; 5.000 – € 7.500; 7.500 – € 10.000; > € 10.000<br>[NL] < € 500; € 500 – € 1.000; € 1.000 – € 2.000; 2.000 – € 3.000; 3.000 – € 4.000; 4.000 – € 5.000; 5.000 – € 7.500; 7.500 – € 10.000; > € 10.000 |
| Sex | [EN] What is your gender? As registered in your passport/ID.<br>[NL] Wat is uw geslacht? Zoals geregistreerd in uw paspoort/identiteitskaart. | [EN] Male; Female; Other, {open text}<br>[NL] Man; Vrouw; Anders, namelijk {open tekst} |
| Gender | [EN] What are your pronouns?<br>[NL] Hoe wordt u het liefst aangesproken? | [EN] He/Him; She/Her; They/Them; Other, namely {open text}<br>[NL] Hij/Hem; Zij/Haar; Die/Hen; Anders, namelijk {open tekst} |
| Ethnicity | [EN] What is your nationality?<br>[NL] Wat is uw nationaliteit? | [EN] Dutch; Belgian; German; Moroccan; Turkish; Surinamese; Other, namely {open text}<br>[NL] Nederlands; Belgisch; Duits; Marokkaans; Turks; Surinaams; Anders, namelijk {open tekst} |
| Nationality at birth | [EN] Is your current nationality different from your nationality at birth?; What was your nationality at birth?<br>[NL] Is uw huidige nationaliteit anders dan bij uw geboorte?; Wat was uw nationaliteit bij geboorte? | [EN] Yes; No; If yes {open text}<br>[NL] Ja; Nee; Als ja {open tekst} |
| Nationality father | [EN] What is the nationality of your father?<br>[NL] Wat is de nationaliteit van uw vader? | [EN] Dutch; Belgian; German; Moroccan; Turkish; Surinamese; Other, namely {open text} |

| | | |
|---|---|---|
| | | [NL] Nederlands; Belgisch; Duits; Marokkaans; Turks; Surinaams; Anders, namelijk {open tekst} |
| Nationality mother | [EN] What is the nationality of your mother?<br>[NL] Wat is de nationaliteit van uw moeder? | [EN] Dutch; Belgian; German; Moroccan; Turkish; Surinamese; Other, namely {open text}<br>[NL] Nederlands; Belgisch; Duits; Marokkaans; Turks; Surinaams; Anders, namelijk |
| Age of onset of overweight and/or obesity | [EN] If you are overweight, at what age did you develop overweight?<br>[NL] Indien u overgewicht heeft, op welke leeftijd kreeg u te maken met overgewicht? | [EN] From (age in years) {numeric}; not applicable<br>[NL] Vanaf (leeftijd in jaren) {numeriek}; niet van toepassing |
| Birth weight | [EN] What was your birth weight? Answer in grams.<br>[NL] Wat was uw gewicht bij geboorte? Geef antwoord in grammen. | [EN] I looked it up {numeric}; This is an estimate {numeric}; I really don't know<br>[NL] Ik heb het opgezocht {numeriek}; Dit is een schatting {numeriek}; Weet ik echt niet |
| Parental BMI (father and mother: current and highest BMI; mother: highest BMI during pregnancy), | [EN] What is your father's age?; What is your father's height?; What is your father's body weight?; What is your mother's age?; What is your mother's height?; What is your mother's body weight?; What was your mother's peak body weight during her pregnancy?<br>[NL] Wat is de leeftijd van uw vader?; Wat is de lengte van uw vader?; Wat is het lichaamsgewicht van uw vader?; Wat is de leeftijd van uw moeder?; Wat is de lengte van uw moeder?; Wat is het lichaamsgewicht van uw moeder?; Wat was het hoogste lichaamsgewicht van uw moeder tijdens haar zwangerschap? | [EN] My father/mother is {numeric, years}; I really don't know / I have no contact with my father/mother / my father/mother is deceased.<br><br>I know for sure, my father/mother is {numeric, cm}; This is an estimate, my father/mother is {numeric, cm}; I really don't know / I have no contact with my father/mother / my father/mother is deceased.<br><br>I know for sure, my father/mother weighs {numeric, kg}; This is an estimate, my father/mother weighs {numeric, kg}; I really don't know / I have no contact with my father/mother / my father/mother is deceased.<br><br>I looked it up/asked, my mother weighed {numeric, kg}; This is an estimate, my mother weighed approximately {numeric, kg}; I really don't know<br>[NL] Mijn vader/moeder is {numeriek, jaren}; Ik weet het echt niet / ik heb geen contact met mijn vader/moeder / mijn vader/moeder is overleden.<br><br>Ik weet het zeker, mijn vader/moeder is {numeriek, cm}; Dit is een schatting, mijn vader/moeder |

| | | |
|---|---|---|
| | | is {numeriek, cm}; Ik weet het echt niet / ik heb geen contact met mijn vader/moeder / mijn vader/moeder is overleden.<br><br>Ik weet het zeker, mijn vader/moeder weegt {numeriek, kg}; Dit is een schatting, mijn vader/moeder weegt {numeriek, kg}; Ik weet het echt niet / ik heb geen contact met mijn vader/moeder / mijn vader/moeder is overleden.<br><br>Ik heb het opgezocht/gevraagd, mijn moeder woog {numeriek, kg}; Dit is een schatting, mijn moeder woog ongeveer {numeriek, kg}; Ik weet het echt niet |
| Smoking status | [EN] Do you smoke nicotine-containing products? (Multiple options possible); How often do you smoke cigarettes/cigars/pipes?; How often do you smoke an e-cigarette or use a nicotine-containing vape?<br>[NL] Rookt u nicotine bevattende middelen? (Meerdere opties mogelijk); Hoe vaak rookt u sigaretten/sigaren/pijp?;<br>Hoe vaak rookt u een e-sigaret of gebruikt u een nicotine-houdende vape? | [EN] No; Yes, cigarettes/cigars/pipe; Yes, vapes/e-cigarettes (not shisha pens, e-hookahs, flavoured vapes)<br><br>Yes, cigarettes/cigars/pipe: Less than once a day/sporadically; <5 times per day; 5–10 times per day; 10–20 times per day; >20 times per day<br><br>Yes, vapes/e-cigarettes: Less than once a day/sporadically; <5 times per day; 5–10 times per day; 10–20 times per day; >20 times per day<br><br>[NL] Nee; Ja, sigaretten/sigaren/pijp; Ja, vape/e-sigaret (niet shisha-pen, e-hookah, vape met smaak)<br><br>Ja, sigaretten/sigaren/pijp: Minder dan een keer per dag/sporadisch; < 5 keer per dag; 5 – 10 keer per dag; 10 – 20 keer per dag; > 20 keer per dag<br><br>Ja, vapes/e-sigaretten: Minder dan een keer per dag/sporadisch; < 5 keer per dag; 5 – 10 keer per dag; 10 – 20 keer per dag; > 20 keer per dag |
| Alcohol and drug use | [EN] How much alcohol do you consume on average per week?; Do you ever use drugs?; | [EN] I don't drink alcohol; Less than 1 drink per week/sporadically; 1 to 7 drinks per week; 8 to 14 drinks |

| | | |
|---|---|---|
| | Which drugs do you use? (Multiple options possible);<br>When did you last use cannabis/hashish?;<br>When did you last use these drugs? (Not cannabis/hashish)<br>[NL] Hoeveel alcohol consumeert u gemiddeld per week?;<br>Gebruikt u wel eens drugs?;<br>Welke drugs gebruikt u? (Meerdere opties mogelijk);<br>Wanneer heeft u voor het laatst cannabis/hasj gebruikt?;<br>Wanneer heeft u voor het laatst deze drugs gebruikt? (Niet cannabis/hasj) | per week; More than 14 drinks per week<br><br>Yes; No<br><br>Cannabis/hashish; MDMA/ecstasy; Ketamine; Psychedelics (magic mushrooms/truffles/LSD); Amphetamine (speed/pep); GHB; Cocaine; Other, namely {open text}<br><br><u>Cannabis/hashish:</u> Past 7 days; Past 30 days; Past 6 months; Past 12 months; More than 12 months ago<br><br><u>Other drugs:</u> Past 30 days; Past 12 months, but not in the past 30 days; More than 12 months ago<br><br>[NL] Ik drink geen alcohol; Minder dan 1 glas per week/sporadisch; 1 tot 7 glazen per week; 8 tot 14 glazen per week; Meer dan 14 glazen per week<br><br>Ja; Nee<br><br>Cannabis/hasj; MDMA/ecstasy; Ketamine; Psychedelica (paddo's/truffels/lsd); Amfetamine (speed/pep); GHB; Cocaïne; Anders, namelijk {open tekst}<br><br><u>Cannabis/hasj:</u> Afgelopen 7 dagen; Afgelopen 30 dagen; Afgelopen 6 maanden; Afgelopen 12 maanden; Meer dan 12 maanden geleden<br><br><u>Andere drugs:</u> Afgelopen 30 dagen; Afgelopen 12 maanden, maar niet in de afgelopen 30 dagen; Meer dan 12 maanden geleden |
| Dietary habits | [EN] Are you currently following a specific diet and/or eating habit?;<br>Have you been eating vegetarian/vegan/pescetarian for more than 3 months?;<br>Have you been following this diet/eating habit for more than 3 months?<br>[NL] Volgt u op dit moment een bepaald dieet en/of eetgewoonte?; | [EN] No; Vegetarian; Pescetarian (no meat, but fish and/or seafood); Vegan/plant-based; Low-carb/low-carb; Paleo; Intermittent fasting; Gluten-free; Raw food; Meal replacements/meal shakes; Other, please specify {open text}<br><u>Vegetarian/Vega/Pescetarian for >3 months:</u> Yes; No<br><u>Other diet >3 months:</u> Yes; No |

| | | |
|---|---|---|
| | Eet u al langer dan 3 maanden vegetarisch/veganistisch/pescotarisch?;<br>Volgt u al langer dan drie maanden dit dieet/deze eetgewoonte? | [NL] Nee; Vegetarisch; Pescotarisch (geen vlees, wel vis en/of zeevruchten); Veganistisch/plantaardig; Koolhydraatbeperkt/koolhydraatarm; Paleo; Intermittent fasting; Glutenvrij; Raw food; Maaltijdvervangers/maaltijdshakes; Anders, namelijk {open tekst}<br>Vegetarisch/vegan/pescotarisch >3 maanden: Ja; Nee<br>Ander dieet >3 maanden: Ja; Nee |
| Contraception use | [EN] Are you using contraception? You can select multiple options: What type of contraception are you using?<br>[NL] Gebruikt u anticonceptie? U kunt meerdere opties kiezen; welke anticonceptie gebruikt u? | [EN] Yes, I use a condom; Yes, I use other contraception; No; I have been sterilized<br><br>Which contraception: Pill; Copper IUD; Hormone IUD; Injection; Hormone rod; Hormone patch; Hormone ring; Other, namely {open text}<br>[NL] Ja, ik gebruik een condoom; Ja, ik gebruik andere anticonceptie; Nee; Ik ben gesteriliseerd<br><br>Welke anticonceptie: Pil; Koperspiraal; Hormoonspiraal; Prikpil; Hormoonstaafje; Hormoonpleister; Hormoonring; Anders, namelijk {open tekst} |
| Menstruation cycle (if applicable) | [EN] When was the first day of your last period?;<br>Do you have regular periods?<br>[NL] Wanneer was de eerste dag van uw laatste ongesteldheid?;<br>Wordt u regelmatig ongesteld?<br>*displayed if gender as in passport is selected as Female | [EN] Enter date [dd/mm/yyyy]; Don't know; I don't have my period (anymore)<br><br>Yes, approximately every 3 weeks; Yes, approximately every 4 weeks; Yes, approximately every 5 weeks; Other, please specify {open text}; No; Don't know<br>[NL] Vul datum in [dd/mm/jjjj]; Weet ik niet; Ik word niet (meer) ongesteld<br><br>Ja, om de 3 weken (ongeveer); Ja, om de 4 weken (ongeveer); Ja, om de 5 weken (ongeveer); Anders, namelijk {open tekst}; Nee; Weet ik niet |
| Medication use | [EN] Are you currently taking any medications? If so, which ones? What are you taking these medications for?<br>[NL] Gebruikt u op dit moment medicijnen? Zo ja, welke?; | [EN] Yes, namely {open text}; No<br>Indication: {open text}<br>[NL] Ja, namelijk {open tekst}; Nee<br>Indicatie: {open tekst} |

| | | |
|---|---|---|
| | Waarvoor gebruikt u deze medicijnen? | |
| Possible other health issues | [EN] Are there any other things we should know about your health?<br>[NL] Zijn er nog andere dingen die wij moeten weten over uw gezondheid? | [EN] Yes, namely {open text}; No<br>[NL] Ja, namelijk {open tekst}; Nee |

#### Beck’s depression inventory II (BDI-II)

| | | | |
|---|---|---|---|
| Reference | Beck, A. T., Steer, R. A., Ball, R., & Ranieri, W. (1996). Comparison of Beck Depression Inventories -IA and -II in psychiatric outpatients. *Journal of personality assessment*, *67*, 588–597. | | |
| Dutch translation | van der Does, A. J. W. (2002). BDI-II-NL. Handleiding. De Nederlandse Versie van de Beck Depression Inventory (2nd ed.). Lisse: Harcourt Test Publishers. | | |
| Total number of items | 21 | Subscales (nr. of items) | 3 subscales: Affect [5 items; 1-2-4-9-12], somatic [9 items; 10-11-15-17-18-19-20-21], cognitive [7 items; 3-5-6-7-8-13-14] |
| Description | The scale consists of 21 statements (e.g., I do not feel sad; I feel sad much of the time) on 3 subscales. Respondents indicate for each statement, which of the 4 responding statements best describes the way they have been feeling over the past 2 weeks about the statement. | | |
| Response format | Respondents select one of the 4 statements per group of statements (indicated with numbers from 0-3).<br>For example:<br>*0 I can concentrate as well as usual*<br>*1 I cannot concentrate as well as usual*<br>*2 It's difficult to keep my mind on something for long.*<br>*3 I can't concentrate well on anything.* | | |
| Skip logic | None. | | |
| Scoring | A total score index is calculated by summing all items (range = 0-63). Items 16 and 18 are a 0-7 scale but are scored 0-3 (statement 1 = 0, 2 and 3 = 1, 4 and 5 = 2, 6 and 7 = 3).<br>Higher scores indicate greater symptom severity<br>Norn scores: minimal: 0-13, light: 14-19, moderately severe: 20-28, severe: 29-63. | | |

#### Power of Food Scale (PFS)

| | |
|---|---|
| Reference | Lowe, M. R., Butryn, M. L., Didie, E. R., Annunziato, R. A., Thomas, J. G., Crerand, C. E., [...], & Halford, J. (2009). The Power of Food Scale. A new measure of the psychological influence of the food environment. *Appetite*, *53*, 114–118. |

| | | | |
|---|---|---|---|
| Dutch translation | Translated by Anita van Oers & Anne Roefs, 2023. Universiteit Maastricht. | | |
| Total number of items | 15 | Subscales (nr. of items) | 3 subscales: food availability [6 items; 1-2-5-10-11-13], presence of food [4 items; 3-4-6-7], food taste [5 items; 8-9-12-14-15] |
| Description | This scale consists of 15 items to assess the psychological impact of living a food-abundant environment. Using 3 subscales; food availability, presence of food, and taste of food, the PFS assess appetite rather than consumption of palatable foods. Respondents indicate for each of the 15 statements to what extent they identify with the statements. Answer options are 1 to 5, from disagree to strongly agree. | | |
| Response format | 1 disagree<br>2 slightly agree<br>3 somewhat agree<br>4 agree<br>5 strongly agree | | |
| Skip logic | None. | | |
| Scoring | A total score of the 15 items is calculated by summing all items (range = 15-75), or total scores per subscale can be calculated. A cut-off is not available, a higher score indicates a greater responsiveness to the food environment. In a sample of individuals with overweight a mean score of 2.57 was found (Cappelleri et al., 2009) | | |

#### Barratt Impulsiveness Scale-11 (BIS-11)

| | | | |
|---|---|---|---|
| Reference | Stanford, M. S., Mathias, C. W., Dougherty, D. M., Lake, S. L., Anderson, N. E., & Patton, J. H. (2009). Fifty years of the Barratt Impulsiveness Scale: An update and review. *Personality and Individual Differences, 47*, 385–395.<br><br>Barratt, E. S. (1994). *Impulsiveness and aggression.* In J. Monahan & H. J. Steadman (Eds.), Violence and mental disorder: Developments in risk assessment (pp. 61–79). The University of Chicago Press. | | |
| Dutch translation | Translated by M. Lijffijt en E. S. Barratt, 2005. | | |
| Total number of items | 30 | Subscales (nr. of items) | 3 subscales: motor [11 items; 2-3-4-16-17-19-21-22-23-25-30], cognitive [8 items; 5-6-9-11-20-24-26-28], non-planning [11 items; 1-7-8-10-12-13-14-15-18-27-29] |
| Description | This questionnaire consists of 30 statements for assessing impulsivity on 3 subscales: motor, cognitive, and non-planning impulsiveness. Respondents indicate on a scale from 1 to 4 how | | |

| | |
|---|---|
| | they felt over the past two weeks, including the day of filling in the questionnaire. |
| Response format | 1 Rarely/never<br>2 Sometimes<br>3 Often<br>4 Always |
| Skip logic | None. |
| Scoring | Items 1-7-8-9-10-12-13-15-20-29-30 are reverse scored (1=4, 2=3, 3=2, and 4=1). A total score is calculated by calculating the sum of all 30 items, or total scores per subscale can be calculated. A cut-off is not available, a higher score indicates a higher degree of impulsiveness. In a female community sample a mean score of 66.41 (sd=11.32) was found (Goodwin, Butler & Nikčević, 2023). |

#### Short Self-Regulation Questionnaire (SSRQ)

| | | | |
|---|---|---|---|
| Reference | Carey, K. B., Neal, D. J., & Collins, S. E. (2004). A psychometric analysis of the self-regulation questionnaire. *Addictive behaviours*, *29*, 253–260.<br>Brown, J. M., Miller, W. R., & Lawendowski, L. A. (1999). The self-regulation questionnaire. | | |
| Dutch translation | Translated by Anita van Oers & Anne Roefs, 2023. Universiteit Maastricht. | | |
| Total number of items | 31 | Subscales (nr. of items) | None |
| Description | This questionnaire consists of 31 statements to measure ability to regulate behaviour in order to achieve desired future outcomes. We use the short form developed by Brown, et al., 1999. Statements include “I doubt I could change even if I wanted to,” “I am able to accomplish goals I set for myself,” “It’s hard for me to notice when I’ve had enough (alcohol, food, sweets),” and “I am able to resist temptation.” These statements are answered by the respondent to what extent they identify with the statements on. Answer options are 1 to 5, from strongly disagree to strongly agree. | | |
| Response format | 1 Strongly disagree<br>2 Disagree<br>3 Uncertain or unsure<br>4 Agree<br>5 Strongly agree | | |
| Skip logic | None. | | |
| Scoring | Items 2-3-4-6-7-9-10-11-16-19-22-23-27 are reverse scored (1=5, 2=4, 3=3, 4=2, and 5=1). A total score is calculated by summing scores on all 31 items for measure of general self-regulation, where lower scores indicate lower self-regulation and higher scores indicate higher self-regulation. Norm scores are not available. | | |

#### Eating Disorder Examination Questionnaire 6.0 (EDE-Q)

<table>
<tr><td>Reference</td><td colspan="3">Berg, K. C., Peterson, C. B., Frazier, P., & Crow, S. J. (2012). Psychometric evaluation of the eating disorder examination and eating disorder examination-questionnaire: a systematic review of the literature. The International journal of eating disorders, 45, 428–438.</td></tr>
<tr><td>Dutch translation</td><td colspan="3">Translated by Anita Jansen, 2014. Universiteit Maastricht. Fairburn, CG & Beglin, SJ (2008). Eating Disorder Examination Questionnaire. In: Fairburn, C (2008). Cognitive Behaviour Therapy and Eating Disorders. Guilford Press. (Unpublished translation by Anita Jansen, Maastricht).</td></tr>
<tr><td>Total number of items</td><td>28</td><td>Subscales (nr. of items)</td><td>4 subscales: restraint [5 items; 1-2-3-4-5], eating concerns [5 items; 7-9-19-20-21], weight concerns [5 items; 8-12-22-24-25], shape concerns [8 items; 6-8-10-11-23-26-27-28], and 6 lose items [binge eating episodes and compensatory behaviour; 13-14-15-16-17-18]</td></tr>
<tr><td>Description</td><td colspan="3">The scale consists of a list of questions (e.g., How dissatisfied have you been with your weight or shape?; Have you had a strong desire to lose weight?).<br>For questions 1-12 and 19-28, respondents indicate how often or to what extent in the last 28 days (4 weeks) the asked behaviour, feelings or cognitions applied to them. For example: “Please indicate: On how many of the past 28 days, did you have a strong desire to lose weight?” or “Has your weight (the number on the scale) influenced how you think (judge) about yourself as a person?”.<br>Lose items; questions 13-18 are answered based on occurrence frequency, e.g. “In the past 28 days, how many times have you eaten what other people would consider an unusually large amount of food (given the circumstances)?”</td></tr>
<tr><td>Response format</td><td colspan="3"><u>Question 1-12</u>:<br>0 No days<br>1 1-5 days<br>2 6-12 days<br>3 13-15 days<br>4 16-22 days<br>5 23-27 days<br>6 Everyday<br><u>Question 13-18</u>:<br>Answer with how many times ‘behaviour’ occurred.<br><u>Question 19-21</u>:<br><u>Q19</u>: answer options as Q1-12<br><u>Q20</u>:<br>0 None of the times<br>1 A few of the times</td></tr>
</table>

| | |
|---|---|
| | 2 Less than half<br>3 Half of the times<br>4 More than half<br>5 Most of the time<br>6 Every time<br>Question 21-28:<br>0 Not at all<br>1 Slightly<br>2 Slightly<br>3 Moderately<br>4 Moderately<br>5 Markedly/Considerable<br>6 Markedly/Considerable |
| Skip logic | None. |
| Scoring | The total score is calculated by adding the calculated subscale scores and dividing this sum score by the number of subscales. For a reliable total score, at least three of the four subscale scores must be calculated. The total score is referred to in English as the "global score."<br>Subscales can be calculated by adding the scores per subscale and dividing by the number of items. For a valid subscore measure at least half of the items needs to be scored ≥1. A higher score indicates higher degree of eating disorder symptoms. In a sample of participants with obesity a global score of 2.75 (sd=0.97) was found (Aardoom, et al., 2012). Considering the subscales, Ivezaj and colleagues (2016) found a mean of 2.1 (sd=1.4) for restraint, 1.3 (sd=1.1) for eating concern, 3.6 (sd=1.5) for shape concern and 3.1 (sd=1.3) for weight concern in sample of participants with overweight and obesity. |

#### Dutch Eating Behaviour Questionnaire (DEBQ)

| | | | |
|---|---|---|---|
| Reference | Van Strien, T., Frijters, J.E.R., Bergers, G.P.A., & Defares, P.B. (1986). The Dutch Eating Behaviour Questionnaire (DEBQ) for assessment of restrained, emotional, and external eating behaviour. International Journal of Eating Disorders, 5(2), 295-315. | | |
| Dutch translation | Van Strien, T., Frijters, J.E.R., Bergers, G.P.A., & Defares, P.B. (1986). The Dutch Eating Behaviour Questionnaire (DEBQ) for assessment of restrained, emotional, and external eating behaviour. International Journal of Eating Disorders, 5(2), 295-315. | | |
| Total number of items | 33 | Subscales (nr. of items) | 3 subscales: emotional eating [13 items; 1-3-5-8-10-13-16-20-23-25-28-30-32], external eating [10 items; 2-6-9-12-15-18-21-24-27-33], dietary restraint [10 items; 4-7-11-14-17-19-22-26-29-31] |
| Description | Questionnaire with 33 statements to assess eating patterns to determine risk factors, individual diagnosis and treatment | | |

| | |
|---|---|
| | strategies for overeating and obesity. Respondents indicate the extent to which each item applies to them, from never to always. Examples of statements are: “When you are irritated, do you feel like eating something?” or “When you see or smell something tasty, do you get hungry?”. |
| Response format | 1 Never<br>2 Seldom<br>3 Sometimes<br>4 Often<br>5 Always |
| Skip logic | None. |
| Scoring | Item 21 is reverse scored (1=5, 2=4, 3=3, 4=2, and 5=1). Scores on all items are summed and divided by 33, or scores per subscale can be calculated by summing per subscale and dividing by number of items. A higher score on emotional eating may indicate an increased risk for obesity or eating disorders, while a high score on external appetite may increase the risk of uncontrolled snacking. Overall, a higher score indicates greater confirmation of disordered eating behaviour. Clinical cut-off scores are not available, and scores should be interpreted on an individual level. Norm scores are available based on a sample of participants with obesity:<br>- Dietary restraint: cut-off M = 2.73<br>- Emotional eating: cut-off M = 2.17<br>- External eating: cut-off M = 2.76<br>(Van Strien, et al., 1986) |

#### Dichotomous Thinking in Eating Disorders Scale (DTEDS-11)

| | | | |
|---|---|---|---|
| Reference | Byrne, S. M., Allen, K. L., Dove, E. R., Watt, F. J., & Nathan, P. R. (2008). *Dichotomous Thinking in Eating Disorders Scale-11 (DTEDS-11)* [Database record]. APA PsycTests. https://doi.org/10.1037/t48296-000 | | |
| Dutch translation | Translated by Anita Jansen. Universiteit Maastricht. | | |
| Total number of items | 11 | Subscales (nr. of items) | 2 subscales:<br>eating [4 items; 1-4-6-8], general [7 items; 2-3-5-7-9-10-11] |
| Description | The scale consists of 11 statements (e.g., I think of food as either “good” or “bad”; Even if I don’t do a job very well, I still think it is worth doing). Respondents indicate the extent to which the statements applied to them over the past month. This scale gives an indication of dichotomous thinking in the context of eating-related problems and is intended for use with dieters, patients with overweight/obesity and patients with eating disorders. | | |
| Response format | 1 Not all true for me<br>2 Slightly true for me<br>3 Fairly true for me<br>4 Very true for me | | |
| Skip logic | None. | | |
| Scoring | An index for each subscale is created by averaging the respective items and a total score is the average of the two subscales scores | | |

| | |
|---|---|
| | (range = 1-4). A higher scores indicates a higher degree of dichotomous thinking. Cut-off scores are not available, Bryne, et al., 2008 find a mean score of 2.61 (sd=0.71) in a sample of patients with obesity. |

#### Body Image Concern Inventory (BICI)

| | | | |
|---|---|---|---|
| Reference | Littleton, H. (2005). *Body Image Concern Inventory (BICI)* [Database record]. APA PsycTests. https://doi.org/10.1037/t00353-000 | | |
| Dutch translation | van Rood, YR. & de Beurs, E. (2005). Nederlandse vertaling van de Body Image Concern Inventory. (H.L. Littleton, , 2005) Vertaald met toestemming van de auteur (H.L. Littleton, , 2005). | | |
| Total number of items | 19 | Subscales (nr. of items) | None. |
| Description | The Body Image Concern Inventory (BICI) is a 19-item self-report measure designed to assess dysmorphic appearance concern. The BICI is used to assess symptom severity. For each item respondents indicated how often they experience the described feeling or perform the described behaviour on a 5-point Likert scale ranging from never to always in the past week. | | |
| Response format | 1 Never<br>2 Seldom<br>3 Sometimes<br>4 Often<br>5 Always | | |
| Skip logic | None. | | |
| Scoring | Scoring ranges from 19 to 95. Different researchers use different subscales (ie. appearance preoccupation, appearance concern, coping behaviour, dysmorphic concern. Nonetheless, all these subscales are highly correlated, thus it recommended to use a single total score. A cut-off-value of 72 has been recommended, such that scores above 72 are regarded as clinical concerning (Shulte-van Maaren, et al., 2014). | | |

#### Rosenberg Self-Esteem Scale-11

| | | | |
|---|---|---|---|
| Reference | Rosenberg, M. (1979). Conceiving the Self. New York: Basic Books | | |
| Dutch translation | Validated Dutch/Flemish translation: Franck, E., De Raedt, R., Barbez, C., & Rosseel, Y. (2008). Psychometric properties of the Dutch Rosenberg self-esteem scale. Psychologica Belgica, 48 (1), 25-35.<br>Dutch translation: Anita Jansen. Universiteit Maastricht. | | |
| Total number of items | 10 | Subscales (nr. of items) | None. |
| Description | The scale consists of a list of statements assessing self-esteem (e.g., At times I think I am no good at all; I take a positive attitude toward myself). Respondents indicate the extent to which the statements are characteristic of them (no time-frame). | | |
| Response format | 0 Strongly disagree<br>1 Disagree | | |

| | |
|---|---|
| | 2 Agree<br>3 Strongly agree |
| Skip logic | None. |
| Scoring | Items 2-5-6-8-9 are reverse scored (strongly agree (0), agree (1), disagree (2), strongly disagree (3)). A total score is calculated as a sum of all items, with a maximum possible score of 30 (range = 0-3). Higher scores indicate a higher level of self-esteem. Average score in Dutch population is 20.9 (sd=4.4) (Franck, et al., 2008), and a score between 15-25 can be seen as a score of normal self-esteem. |

#### Behavioural Regulation of Exercise Questionnaire (BREQ-2) & Regulation of Eating Behaviour Scale (REBS)

| | | | |
|---|---|---|---|
| Reference(s) | Markland, D., & Tobin, V. (2004). A modification to the behavioural regulation in exercise questionnaire to include an assessment of amotivation. *Journal of Sport and Exercise Psychology*, *26*(2), 191-196.<br>Pelletier, L. G., Dion, S. C., Slovinec-D'Angelo, M., & Reid, R. (2004). Why do you regulate what you eat? Relationships between forms of regulation, eating behaviours, sustained dietary behaviour change, and psychological adjustment. *Motivation and emotion*, *28*(3), 245-27 | | |
| Dutch translation | Helmink, J.H.M., Van Boekel, L.C., van der Sluis, M.E. & Kremers, S.P.J. (2011). Lange termijn evaluatie onder deelnemers aan de BeweegKuur: Rapportage van de resultaten van een follow-up meting bij deelnemers. Universiteit Maastricht | | |
| Total number of items | 24 (1-12 items for physical activity, 13-24 items for healthy eating) | Subscales (nr. of items) | 5 subscales: amotivation [4 items; 1-2-13-14], external regulation [4 items; 3-4-15-16], introjected regulation [4 items; 5-6-17-18], identified regulation [6 items; 7-8-9-19-20-21], and intrinsic motivation [6 items; 10-11-12-22-23-24] |
| Description | The BREQ-2 focuses on reasons why people exercise, and the REBS focuses on reasons why people eat healthy. For both, we used the shortened 12-item questionnaire. Both questionnaires are based on the self-determination theory and includes statements on amotivation, external regulation, introjected regulation, identified regulation and intrinsic motivation to measure the continuum of behavioural regulation in the context of exercise and healthy eating.<br>Some examples of statements for the BREQ-2 are ‘I exercise because other people say I should’, or ‘I feel ashamed when I miss an exercise session’. For the REBS some examples are ‘I eat healthy …’; ‘Because it is expected of me’, ‘Because eating healthy is an integral part of my lifestyle’, or ‘I don’t know. I can’t see how | | |

| | |
|---|---|
| | my efforts to eat healthy are helping my health situation'. All these statements are scored on a scale from 0 to 4, from strongly disagree to strongly agree. |
| Response format | 0 Strongly disagree<br>1 Disagree<br>2 Not agree/not disagree<br>3 Agree<br>4 Strongly agree |
| Skip logic | None. |
| Scoring | A total score or relative autonomous index (RAI) is calculated by weighting each subscale and summing the weighted scores: (amotivation multiplied by -3) + (external regulation multiplied by -2) + (introjected regulation multiplied by -1) + (identified regulation multiplied by 2) + (intrinsic regulation multiplied by 3). Scores per subscale are added up to a total score of physical activity (BREQ) or healthy eating (REBS). Each of these subscales is weighted and summed, to calculate an RAI, adding up to a total score between -24 and +20, where higher scores indicate more autonomous motivation and lower scores indicated more self-controlled regulation of amotivation. |

#### Health Status/QoL - Euroqol-5

| | | | |
|---|---|---|---|
| Reference | EuroQoL Group 1990 | | |
| Dutch translation | EuroQoL Group 1990 | | |
| Total number of items | 6 | Subscales (nr. of items) | 6 subscales: mobility, self-care, activities of daily living, pain/complaints, mood, overall well-being |
| Description | The Euro-Quality of Life (QoL) is a standardized, non-disease specific instrument for describing and valuing health states. It is intended to complement other quality-of-life measures and to facilitate the collection of a common data set for reference purposes. The EuroQoL includes five statements on mobility, self-care, activities of daily living, pain/complaints and mood. These statements are rated from no problems with this activity (1) to severe problems (3). Item 6 asks about general health status the day of filling in the questionnaire on a scale from 0 to 100, as a utility score of quality of life (from poor to good). | | |
| Response format | <u>Item 1 to 5</u>:<br>1 No problems<br>2 Some problems<br>3 Severe problems<br><u>Item 6</u>:<br>Scale of 0-100 for health; 0 = worst imaginable health status, 100 = best imaginable health status | | |
| Skip logic | None. | | |
| Scoring | A sum score is calculated for items 1-5 (range 5-15), a higher score indicates more restrictions. Item 6 is evaluated separately and can be analysed on a population/study sample level. | | |

#### Home Address

| Variable | Item | Response format |
|---|---|---|
| [EN] What is your postal code?<br>[NL] Wat is uw postcode? | [EN] Postal code<br>[NL] Post code | [EN] 6 digit postal code<br>[NL] 6-cijferige postcode |
| [EN] What is your street name and house number?<br>[NL] Wat is uw straatnaam en huisnummer? | [EN] Street name and house number<br>[NL] Straatnaam en huisnummer | [EN] Street name + house number<br>[NL] Straatnaam + huisnummer |
| Description | This data is asked to collect information on a participant's physical environment. Based on postal code we can collect information on availability of supermarkets, fast-food chains, restaurants, green spaces, and sports facilities in someone's neighbourhood. Furthermore, the safety of a neighbourhood can be evaluated, as this might impact if someone will walk somewhere, or rather bike or drive., This is also influenced by walkability, availability of bike paths, availability of public transport or connectedness of roads. We aim to determine if a participant's physical environment contributes positively or negatively to their lifestyle. | |

#### Social Support for Eating Habits + Social Support for Exercise Survey

| | | | |
|---|---|---|---|
| Reference | Sallis, J.F., Grossman, R.M., Pinski, R.B., Patterson, T.L., and Nader, P.R. (1987). The development of scales to measure social support for diet and exercise behaviours. Preventive Medicine, 16, 825-836. | | |
| Dutch translation | Translated by Anita van Oers & Anne Roefs, 2023. Universiteit Maastricht. | | |
| Total number of items | 46, 23 for family and 23 for friends | Subscales (nr. of items) | 2 subscales: healthy eating [10 items; 1-10, 24-33], exercise [13 items; 11-23, 34-46]. Additional subscales: encouragement eating [5 items: 1-5], discouragement [5 items: 6-10], participation exercise [10 items: 11-16, 20-23] |
| Description | This questionnaire includes a total of 46 statements, 23 statements for support from family and the same 23 statements for support from friends. These statements are measures of perceived social support specific to health-related eating and exercise behaviour. Each question is thus answered twice, and respondents indicate how often relatives or household members, friends, colleagues, or … have said or done the described item in the past three months. Examples of statements are; "...encouraged me not to eat | | |

| | |
|---|---|
| | "unhealthy foods" (cake, salted chips) when I was tempted." or "...complained about the time I spend on sports.". |
| Response format | 1 Never<br>2 Seldom<br>3 A few times<br>4 Often<br>5 Very often<br>8 Not applicable |
| Skip logic | None. |
| Scoring | Sum scores can be calculated per subscale, separately for relatives (or household members) and friends. Scores of "8" are recoded to "1". A higher score indicates higher support for healthy eating or exercise. |

#### Exercise Vital Sign Questionnaire

| | | | |
|---|---|---|---|
| Reference | Coleman, K. J., Ngor, E., Reynolds, K., Quinn, V. P., Koebnick, C., Young, D. R., Sternfeld, B., & Sallis, R. E. (2012). Initial validation of an exercise "vital sign" in electronic medical records. Medicine and science in sports and exercise, 44, 2071–2076. | | |
| Dutch translation | Translated by Anita van Oers & Anne Roefs, 2023. Universiteit Maastricht. | | |
| Total number of items | 3 | Subscales (nr. of items) | None. |
| Description | The Exercise Vital Sign (EVS) Questionnaire is a brief physical activity questionnaire to measure physical activity as a vital sign and to determine if a participant meets the current physical recommendations. | | |
| Response format | Q1 Days per week of moderate to vigorous activity.<br>Q2 Minutes activity at this level (moderate to vigorous) per day.<br>Q3 Days per week of muscle strengthening activities. | | |
| Skip logic | None. | | |
| Scoring | Question 1 and 2 are multiplied for minutes of physical activity per week. Question 3 is scored separately. Both can be compared to the Dutch recommended physical activity guidelines. Current guidelines are a minimum of 150 minutes of moderate to vigorous intensity exercise, and muscle strengthening activities at least twice a week (Kenniscentrum Sport & Bewegen, beweegrichtlijnen). | | |

#### PrimeScreen

| | | | |
|---|---|---|---|
| Reference | Rifas-Shiman, S. L., Willett, W. C., Lobb, R., Kotch, J., Dart, C., & Gillman, M. W. (2001). PrimeScreen, a brief dietary screening tool: reproducibility and comparability with both a longer food frequency questionnaire and biomarkers. *Public Health Nutrition*, *4*(2), 249–254. doi:10.1079/PHN200061 | | |
| Dutch translation | Translated by Anita van Oers & Anne Roefs, 2023. Universiteit Maastricht. | | |
| Total number of items | 21 | Subscales (nr. of items) | None. |

| | |
|---|---|
| Description | The PrimeScreen questionnaire is a general dietary screening tool to assess known relationships between dietary factors and major causes of morbidity and mortality. The original questionnaire includes 18 items about the average frequency of consumption, over the previous year, of specified foods and food groups, and another seven items about vitamin and supplement intake. All 18 items include examples of the most frequently consumed foods that constitute that group. PrimeScreen particularly targets intake of fruits, vegetables, whole and low-fat dairy products, whole grains, fish and red meat as well as other foods that are major contributors to the intake of saturated and trans-fat. Three items were added to also be inclusive for a vegetarian or vegan diet as this is common in the Netherlands. As the original questionnaire consists of 18 items, a total of 21 items was used. All items are scored from less than once a week to twice or more per day.<br><br>Added items:<br>[NL] Plantaardige zuivelalternatieven (sojayoghurt, kokosmelk, haverdrank) / [EN] Plant-based dairy alternatives (soy yogurt, coconut milk, oat drink)<br>[NL] Peulvruchten (linzen, sojabonen, kikkererwten) / [EN] Legumes (lentils, soybeans, chickpeas)<br>[NL] Vleesvervangers (tofu, tempeh, vega burger) / [EN] Meat substitutes (tofu, tempeh, veggie burger) |
| Response format | Less than once a week<br>Once a week<br>2-4 times a week<br>Almost daily or daily<br>Twice or more a day |
| Skip logic | None. |
| Scoring | The Primescreen survey is scored using the Traffic light method (Kronsteiner-Gicevic, et al., 2023). Participants would receive 2 points (green) if they ate the product frequently and in accordance with Dutch dietary guidelines (Voedingscentrum), 1 point if the food was consumed frequently (orange), but consumption could be improved. And 0 points if the food was not consumed frequently enough (red). Unhealthy items are reverse scored. |

#### Questionnaire for family and friends

##### *General information*

| Variable | Item | Response format |
|---|---|---|
| Relationship to subject | [EN] What is your relationship to the subject?<br>[NL] Wat is uw relatie tot de proefpersoon? | [EN] Partner, family, friend, neighbours, other [text entry]<br>[NL] Partner, familie, vriend(in), buren, anders [tekst entry] |
| Age | [EN] What is your age?<br>[NL] Wat is uw leeftijd? | [EN] Age in years {numeric}<br>[NL] Leeftijd in jaren |
| Current weight | [EN] What is your current weight?<br>[NL] Wat is uw huidig gewicht? | [EN] Weight in kg {numeric}<br>[NL] Gewicht in kg {numeriek} |

| | | |
|---|---|---|
| Length | [EN] What is your length?<br>[NL] Wat is uw lengte? | [EN] Length in cm {numeric}<br>[NL] Lengte in cm {numeriek} |

*International Physical Activity Questionnaire (IPAQ)*

| | | | |
|---|---|---|---|
| Reference | IPAQ, 1998 | | |
| Dutch translation | Translated by Jikke Hesen & Eva Vanbrabant, 2023. Universiteit Maastricht. | | |
| Total number of items | 31 items | Subscales (nr. of items) | 5 subscales: physical activities at work [8 items]; physical activities related to transportation [8 items]; housework, chores, and family responsibilities [6 items]; physical activities related to sports, recreation, and leisure [7 items]; time spent sitting [2 items]. |
| Description | The International Physical Activity Questionnaire (IPAQ) is used to assess physical activity levels across 5 different sub-areas. For all sub-areas it asks about days per week, time per day (in minutes), and considers different intensities. For example: "On how many days in the last seven days did you walk for at least 10 minutes at a time as part of your work?" answering with 0 to 7 days a week, followed by "How much time in total did you spend walking on such day(s) as part of your work? Answer in minutes per day.", followed by "If you walked as part of your work, what was your typical pace? Did you walk in:" answering with low pace, moderate pace or fast pace. | | |
| Response format | Days per week; followed by minutes per day; followed by tempo/intensity of activity (e.g., Walking at low, moderate or high pace) | | |
| Skip logic | None. | | |
| Scoring | Count number of days, or minutes per day, intensity of activity per subscale or question. A higher score corresponds to a more physically demanding activity. The IPAQ can be scored as a categorical variable with the intensity level of an activity (low, moderate or high) or as a continuous variable with minutes of the activity per week.<br><br>Physical activities are scored as follows to include moderate-intensity activities between 3 and 6 METs and vigorous-intensity activities as >6 METs. The weighted MET-minutes per week (MET·min·wk) were calculated as duration × frequency per week × MET intensity. These are summed across activity domains to produce a weighted estimate of total physical activity from all reported activities per week (MET·min·wk). (see Craig, et al., 2003; DOI: 10.1249/01.MSS.0000078924.61453.FB) | | |

### Ecological Momentary Assessment (EMA)

#### Lifestyle survey

*this ‘lifestyle survey’ triggered semi-randomized 8 times per day as part of the baseline and post-intervention (duplicates) 3-week EMA measurement period.

| Variable | Item | Answer options |
|---|---|---|
| [EN] Total positive affect<br>[NL] Totaal positive affect | [EN] How strong are your positive feelings right now?<br>[NL] Hoe sterk zijn uw positieve gevoelens op dit moment? | [EN] VAS scale 0-100: ‘Not at all strong’ to ‘very strong’<br>[NL] VAS schaal 0-100: ‘Helemaal niet sterk’ tot ‘heel erg sterk’ |
| [EN] Total negative affect<br>[NL] Totaal negatieve affect | [EN] How strong are your negative feelings right now?<br>[NL] Hoe sterk zijn uw negative gevoelens op dit moment? | [EN] VAS scale 0-100: ‘Not at all strong’ to ‘very strong’<br>[NL] VAS schaal 0-100: ‘Helemaal niet sterk’ tot ‘heel erg sterk’ |
| [EN] Scared/anxious<br>[NL] Bang/angstig | [EN] How scared/anxious do you feel right now?<br>[NL] Hoe bang/angstig voelt u zich op dit moment? | [EN] VAS scale 0-100: ‘Not at all scared/anxious’ to ‘very afraid/anxious’<br>[NL] VAS schaal 0-100: ‘Helemaal niet bang/angstig’ tot ‘heel erg bang/angstig’ |
| [EN] Irritated/angry<br>[NL] Geïrriteerd/boos | [EN] How irritated/angry do you feel right now?<br>[NL] Hoe geïrriteerd/boos voelt u zich op dit moment? | [EN] VAS scale 0-100: ‘Not at all irritated/angry’ to ‘very irritated/angry’<br>[NL] VAS schaal 0-100: ‘Helemaal niet geïrriteerd/boos’ tot ‘heel erg geïrriteerd/bos’ |
| [EN] Stress<br>[NL] Stress | [EN] How stressed are you right now?<br>[NL] Hoe gestrest bent u op dit moment? | [EN] VAS scale 0-100: ‘Not at all stressed’ to ‘very stressed’<br>[NL] VAS schaal 0-100: ‘Helemaal niet gestrest’ tot ‘heel erg gestrest’ |
| [EN] Happy/cheerful<br>[NL] Blij/vrolijk | [EN] How happy/cheerful do you feel right now?<br>[NL] Hoe blij/vrolijk voelt u zich op dit moment? | [EN] VAS scale 0-100: ‘Not at all happy/cheerful’ to ‘very happy/cheerful’<br>[NL] VAS schaal 0-100: Helemaal niet blij/vrolijk tot heel erg blij/vrolijk |
| [EN] Sad/gloomy<br>[NL] Droevig/somber | [EN] How sad/gloomy do you feel right now?<br>[NL] Hoe droevig/somber voelt u zich op dit moment? | [EN] VAS scale 0-100: ‘Not at all sad/gloomy’ to ‘very sad/gloomy’<br>[NL] VAS schaal 0-100: ‘Helemaal niet droevig/somber’ tot ‘heel erg droevig/somber’ |

| | | |
|---|---|---|
| [EN] Bored<br>[NL] Verveeld | [EN] How bored are you right now?<br>[NL] Hoe verveeld voelt u zich op dit moment? | [EN] VAS scale 0-100: 'Not at all bored' to 'very bored'<br>[NL] VAS schaal 0-100: 'Helemaal niet verveeld' tot 'heel erg verveeld' |
| [EN] Energetic<br>[NL] Energiek | [EN] How energetic do you feel right now?<br>[NL] Hoe energiek voelt u zich op dit moment? | [EN] VAS scale 0-100: 'Not at all energetic' to 'very energetic'<br>[NL] VAS schaal 0-100: 'Helemaal niet energiek' tot 'heel erg energiek' |
| [EN] Confident<br>[NL] Zelfverzekerd | [EN] How confident do you feel right now?<br>[NL] Hoe zelfverzekerd voelt u zich op dit moment? | [EN] VAS scale 0-100: 'Not at all confident' to 'very confident'<br>[NL] VAS schaal 0-100: 'Helemaal niet zelfverzekerd' tot 'heel erg zelfverzekerd' |
| [EN] Ashamed/guilty<br>[NL] Beschaamd/schuldig | [EN] How ashamed/guilty do you feel right now?<br>[NL] Hoe beschaamd/schuldig voelt u zich op dit moment? | [EN] VAS scale 0-100: 'Not at all ashamed/guilty' to 'very ashamed/guilty'<br>[NL] VAS schaal 0-100: 'Helemaal niet beschaamd/schuldig' tot 'heel erg beschaamd/schuldig' |
| [EN] Lonely<br>[NL] Eenzaam | [EN] How lonely do you feel right now?<br>[NL] Hoe eenzaam voelt u zich op dit moment? | [EN] VAS scale 0-100: 'Not at all lonely' to 'very lonely'<br>[NL] VAS schaal 0-100: 'Helemaal niet eenzaam' tot 'heel erg eenzaam' |
| [EN] Context: Who<br>[NL] Context: Wie | [EN] Who are you with right now? You can select multiple answers.<br>[NL] Met wie bent u op dit moment? U kunt meerdere antwoorden selecteren. | [EN] No one; Partner; Friends; Family members/housemates; Other family members; Colleagues/classmates; Strangers; Other {open text}<br>[NL] Niemand; Partner; Vrienden; Gezinsleden/huisgenoten; Overige familieleden; Collega's/klasgenoten; Onbekenden; Anders {open tekst} |

| | | |
|---|---|---|
| [EN] Context: Where<br>[NL] Context: Waar | [EN] Where are you right now?<br>[NL] Waar bent u op dit moment? | [EN] At home; Guest at someone's house; Work/school; Restaurant/café; On the road/on the go; Shop/supermarket; Gym/(sports)club; Outdoors; Other {open text}<br>[NL] Thuis; Te gast bij iemand; Werk/school; Restaurant/café; Onderweg; Winkel/supermarkt; Sportschool/vereniging/club; Buiten; Anders {open tekst} |
| [EN] Social media<br>[NL] Sociale media | [EN] How much time did you spend on social media in the past 2 hours?<br>[NL] Hoeveel tijd heeft u in de afgelopen 2 uur aan social media besteed? | [EN] 0 minutes; Less than 15 minutes; 15 to 30 minutes; 30 minutes to 1 hour; More than 1 hour.<br>[NL] 0 minuten; Minder dan 15 minuten; 15 tot 30 minuten; 30 minuten tot 1 uur; Meer dan 1 uur. |
| [EN] Physical activity duration<br>[NL] Fysieke activiteit duur | [EN] How long were you physically active in the past 2 hours?<br>[NL] Hoe lang bent u in de afgelopen 2 uur fysiek actief geweest? | [EN] 0 minutes; Less than 15 minutes; 15 to 30 minutes; 30 minutes to 1 hour; 1 hour to 1.5 hours; More than 1.5 hours.<br>[NL] 0 minuten; Minder dan 15 minuten; 15 tot 30 minuten; 30 minuten tot 1 uur; 1 uur tot 1.5 uur; Meer dan 1.5 uur. |
| [EN] Physical activity type<br>[NL] Fysieke activiteit soort | [EN] What did you do? You can select multiple answers.<br>[NL] Wat heeft u gedaan? U kunt meerdere antwoorden selecteren. | [EN] Walking; Running; Cycling; Strength training; Cardio training; Gardening; Cleaning; Swimming; Ball sports/team sports; Other {open text}<br>[NL] Wandelen; Hardlopen; Fietsen; Krachttraining; Cardiotraining; Tuinieren; Schoonmaken; Zwemmen; Balsport/teamsport; Anders {open tekst} |
| [EN] Physical activity intensity walking<br>[NL] Fysieke activiteit intensiteit wandelen | [EN] How intensive was your walk?<br>[NL] Hoe intensief heeft u gewandeld? | [EN] VAS scale 0-100: 'Light intensive' to 'heavy intensive'<br>[NL] VAS schaal 0-100: 'Licht intensief' tot 'zwaar intensief' |

| | | |
|---|---|---|
| [EN] Physical activity intensity running<br>[NL] Fysieke activiteit intensiteit hardlopen | [EN] How intense was your run?<br>[NL] Hoe intensief heeft u hardgelopen? | [EN] VAS scale 0-100: ‘Light intensive’ to ‘heavy intensive’<br>[NL] VAS schaal 0-100: ‘Licht intensief’ tot ‘zwaar intensief’ |
| [EN] Physical activity intensity cycling<br>[NL] Fysieke activiteit intensiteit fietsen | [EN] How intensive was the cycling?<br>[NL] Hoe intensief heeft u gefietst? | [EN] VAS scale 0-100: ‘Light intensive’ to ‘heavy intensive’<br>[NL] VAS schaal 0-100: ‘Licht intensief’ tot ‘zwaar intensief’ |
| [EN] Physical activity intensity strength training<br>[NL] Fysieke activiteit intensiteit krachttraining | [EN] How intense was your strength training?<br>[NL] Hoe intensief was uw krachttraining? | [EN] VAS scale 0-100: ‘Light intensive’ to ‘heavy intensive’<br>[NL] VAS schaal 0-100: ‘Licht intensief’ tot ‘zwaar intensief’ |
| [EN] Physical activity intensity cardio training<br>[NL] Fysieke activiteit intensiteit cardiotraining | [EN] How intense was your cardio workout?<br>[NL] Hoe intensief was uw cardiotraining? | [EN] VAS scale 0-100: ‘Light intensive’ to ‘heavy intensive’<br>[NL] VAS schaal 0-100: ‘Licht intensief’ tot ‘zwaar intensief’ |
| [EN] Physical activity intensity gardening<br>[NL] Fysieke activiteit intensiteit tuinieren | [EN] How intensive was the gardening?<br>[NL] Hoe intensief heeft u getuinierd? | [EN] VAS scale 0-100: ‘Light intensive’ to ‘heavy intensive’<br>[NL] VAS schaal 0-100: ‘Licht intensief’ tot ‘zwaar intensief’ |
| [EN] Physical activity intensity cleaning<br>[NL] Fysieke activiteit intensiteit schoonmaken | [EN] How intensive was the cleaning?<br>[NL] Hoe intensief heeft u schoongemaakt? | [EN] VAS scale 0-100: ‘Light intensive’ to ‘heavy intensive’<br>[NL] VAS schaal 0-100: ‘Licht intensief’ tot ‘zwaar intensief’ |
| [EN] Physical activity intensity swimming<br>[NL] Fysieke activiteit intensiteit zwemmen | [EN] How intensive was the swimming?<br>[NL] Hoe intensief heeft u gezwommen? | [EN] VAS scale 0-100: ‘Light intensive’ to ‘heavy intensive’<br>[NL] VAS schaal 0-100: ‘Licht intensief’ tot ‘zwaar intensief’ |
| [EN] Physical activity intensity ball sport/team sport<br>[NL] Fysieke activiteit intensiteit balsport/teamsport | [EN] How intensive was the ball sport/team sport?<br>[NL] Hoe intensief was uw balsport/teamsport? | [EN] VAS scale 0-100: ‘Light intensive’ to ‘heavy intensive’<br>[NL] VAS schaal 0-100: ‘Licht intensief’ tot ‘zwaar intensief’ |
| [EN] Physical activity intensity other<br>[NL] Fysieke activiteit intensiteit anders | [EN] How intensive was the other physical activity?<br>[NL] Hoe intensief was uw fysieke activiteit? | [EN] VAS scale 0-100: ‘Light intensive’ to ‘heavy intensive’<br>[NL] VAS schaal 0-100: ‘Licht intensief’ tot ‘zwaar intensief’ |

| | | |
|---|---|---|
| [EN] Craving<br>[NL] Trek | [EN] How much are you craving food right now?<br>[NL] Hoeveel trek in eten heeft u op dit moment? | [EN] VAS scale 0-100: ‘No cravings at all’ to ‘irresistible cravings’<br>[NL] VAS schaal 0-100: ‘Helemaal geen trek’ tot ‘onweerstaanbare trek’ |
| [EN] Specific craving<br>[NL] Specifieke trek | [EN] Are you craving something specific?<br>[NL] Heeft u trek in iets specifieks? | [EN] Yes; No<br>[NL] Ja; Nee |
| [EN] Specific craving category<br>[NL] Specifieke trek categorie | [EN] What are you craving? You can select multiple answers.<br>[NL] Waar heeft u trek in? U kunt meerdere antwoorden selecteren. | [EN] Bread/cereal/pancakes; Dairy/egg; Vegetables/fruit; Soup; Meal; Snack; Pastry/dessert<br>[NL] Brood/ontbijtgranen/pannen koeken; Zuivel/ei; Groente/fruit; Soep; Maaltijd; Tussendoortje/snack; Gebak/dessert |
| [EN] Eating moments<br>[NL] Eetmomenten<br>(ie. an eating moment is any moment someone eats anything, this can be a snack or a meal) | [EN] How many eating moments have you had since you last completed the questionnaire?<br>[NL] Hoeveel eetmomenten heeft u sinds de laatste ingevulde vragenlijst gehad? | [EN] None; 1 eating moment; 2 eating moments; 3 eating moments; 4 eating moments; 5 eating moments<br>[NL] Geen; 1 eetmoment; 2 eetmomenten; 3 eetmomenten; 4 eetmomenten; 5 eetmomenten<br>**the following are repeated for the amount eating moments indicated* |
| *[EN] Food category<br>[NL] Eetcategorie | [EN] What did you eat? You can select multiple answers.<br>[NL] Wat heeft u gegeten? U kunt meerdere antwoorden selecteren. | [EN] Bread/cereal/pancakes; Dairy/egg; Vegetables/fruit; Soup; Meal; Snack; Pastry/dessert<br>[NL] Brood/ontbijtgranen/pannen koeken; Zuivel/ei; Groente/fruit; Soep; Maaltijd; Tussendoortje/snack; Gebak/dessert |

| | | |
|---|---|---|
| *[EN] Subcategory: Bread/cereals/pancakes<br>[NL] Subcategorie: Brood/ontbijtgranen/pannenkoeken | [EN] Bread/cereal/pancakes: what did you eat? You can select multiple answers.<br>[NL] Brood/ontbijtgranen/pannenkoeken: wat heeft u gegeten? U kunt meerdere antwoorden selecteren. | [EN] Crackers/rusks/rice wafers (with toppings); Baguette/sandwich (with toppings); Sweet rolls/pancakes/little Dutch pancakes/French toast; Porridge/oatmeal/muesli (with/without milk); Cruesli/granola (with/without milk); Cornflakes/sweet breakfast cereals (with/without milk)<br>[NL] Cracker/beschuit/rijstwafel (met beleg); (Stok)brood(je)/boterham (met beleg); Zoet brood(je)/pannenkoeken/poffertjes/wentelteefjes; Ontbijtpap/havermout/muesli (met/zonder melk); Cruesli/granola (met/zonder melk); Cornflakes/zoete ontbijtgranen (met/zonder melk) |
| *[EN] Subcategory: dairy/egg<br>[NL] Subcategorie: zuivel/ei | [EN] Dairy/eggs: What did you eat? You can select multiple answers.<br>[NL] Zuivel/ei: wat heeft u gegeten? U kunt meerdere antwoorden selecteren. | [EN] (Plant-based) yogurt/quark; Egg<br>[NL] (Plantaardige) yoghurt/kwark; Ei |
| *[EN] Subcategory: vegetables/fruit<br>[NL] Subcategorie: groente/fruit | [EN] Vegetables/fruit: what did you eat? You can select multiple answers.<br>[NL] Groente/fruit: wat heeft u gegeten? U kunt meerdere antwoorden selecteren. | [EN] Fruit/fruit salad; (snack/small) vegetables; Side salad; Dried fruit<br>[NL] Fruit/fruitsalade; (Snoep/snack)groente; Bijgerecht salade; Gedroogde vruchten |
| *[EN] Subcategory: Soup<br>[NL] Subcategorie: Soep | [EN] Soup: What did you eat? You can select multiple answers.<br>[NL] Soep: wat heeft u gegeten? U kunt meerdere antwoorden selecteren. | [EN] Soup; Filled soup/meal soup<br>[NL] Soep; Gevulde soep/maaltijdsoep |

| | | |
|---|---|---|
| *[EN] Subcategory: Meal<br>[NL] Subcategorie: Maaltijd | [EN] Meal: What did you eat? You can select multiple answers.<br>[NL] Maaltijd: wat heeft u gegeten? U kunt meerdere antwoorden selecteren. | [EN] Potato-vegetable-meat/fish/meat substitute; Rice/noodle/pasta dish; Couscous/bulgur/quinoa dish; Filled roll/wrap/taco/pita; Quiche/savoury pie/homemade pizza; Sushi/tapas; fries/fried snack/pizza; meal salad<br>[NL] Aardappel-groente-vlees/vis/vleesvervanger; Rijst/noedel/pasta-gerecht; Couscous/bulgur/quinoa-gerecht; Gevuld broodje/wrap/taco/pita; Quiche/hartige taart/zelfgemaakte pizza; Sushi/tapas; friet/gefrituurde snack/pizza; maaltijdsalade |
| *[EN] Subcategory: snack<br>[NL] Subcategorie: tussendoortje/snack | [EN] Snack: What did you eat? You can select multiple answers.<br>[NL] Tussendoortje/snack: wat heeft u gegeten? U kunt meerdere antwoorden selecteren. | [EN] Cereal bar/protein bar/breakfast biscuit/fruit biscuit; Cookie/waffle; Chips/salted toast or nuts/popcorn; Chocolate (bar)/M&Ms/candy; Unsalted nuts/kernels/seeds; Fried snack; charcuterie<br>[NL] Granenreep/proteïnereep/ontbijtkoek/fruitbiscuit; Koekje/wafel; Chips/gezouten toastjes of noten/popcorn; Chocolade(reep)/M&Ms/snoep; Ongezouten noten/pitten/zaden; Gefrituurde snack; kaasjes/worstjes |
| *[EN] Subcategory: Pastry/Dessert<br>[NL] Subcategorie: Gebak/dessert | [EN] Pastry/dessert: What did you eat? You can select multiple answers.<br>[NL] Gebak/dessert: wat heeft u gegeten? U kunt meerdere antwoorden selecteren. | [EN] Cake/pastry/pie/candies/profiteroles; Ice cream/ice cream cake; Custard/pudding/dessert; Water ice/sorbet; Cheese board<br>[NL] Taart/gebak/vlaai/bonbons/soesjes; Roomijs/ijstaart; Vla/pudding/dessert; Waterijs/sorbetijs; Kaasplank |

| | | |
|---|---|---|
| *[EN] Mealtime time of day<br>[NL] Eetmoment tijd | [EN] What time approximately did you eat this?<br>[NL] Hoe laat heeft u dit ongeveer gegeten? | [EN] Between 6:00 and 8:00; Between 8:00 and 10:00; Between 10:00 and 12:00; Between 12:00 and 14:00; Between 14:00 and 16:00; Between 16:00 and 18:00; Between 18:00 and 20:00; Between 20:00 and 22:00<br>[NL] Tussen 06:00 en 08:00; Tussen 08:00 en 10:00; Tussen 10:00 en 12:00; Tussen 12:00 en 14:00; Tussen 14:00 en 16:00; Tussen 16:00 en 18:00; Tussen 18:00 en 20:00; Tussen 20:00 en 22:00 |
| *[EN] Mealtime plan<br>[NL] Eetmoment plan | [EN] At this mealtime I…<br>[NL] Op dit eetmoment heb ik… | [EN] Ate much less than I planned; Ate less than I planned; Ate exactly what I planned; Ate more than I planned; Ate much more than I planned<br>[NL] Veel minder gegeten dan ik van plan was; Minder gegeten dan ik van plan was; Precies gegeten wat ik van plan was; Meer gegeten dan ik van plan was; Veel meer gegeten dan ik van plan was |
| *[EN] Mealtime emotion<br>[NL] Eetmoment emotie | [EN] How did you feel when you ate this? You can select up to two answers.<br>[NL] Hoe voelde u zich toen u dit at? U kunt maximaal 2 antwoorden selecteren. | [EN] Scared/anxious; Irritated/angry; Stressed; Relaxed/calm; Happy/cheerful; Sad/gloomy; Bored; Tired; Energetic; Confident; Insecure; Ashamed/guilty; Lonely<br>[NL] Bang/angstig; Geïrriteerd/boos; Gestresst; Ontspannen/kalm; Blij/vrolijk; Droevig/somber; Verveeld; Vermoeid; Energiek; Zelfverzekerd; Onzeker; Beschaamd/schuldig; Eenzaam |

| | | |
|---|---|---|
| *[EN] Mealtime intensity scared<br>[NL] Eetmoment intensiteit bang | [EN] How scared/anxious did you feel during this meal?<br>[NL] Hoe bang/angstig voelde u zich tijdens dit eetmoment? | [EN] VAS scale 0-100: ‘Not at all scared/anxious’ to ‘very afraid/anxious’<br>[NL] VAS schaal 0-100: ‘Helemaal niet bang/angstig’ tot’ heel erg bang/angstig’ |
| *[EN] Mealtime intensity irritated<br>[NL] Eetmoment intensiteit geïrriteerd | [EN] How irritated/angry did you feel during this meal?<br>[NL] Hoe geïrriteerd/boos voelde u zich tijdens dit eetmoment? | [EN] VAS scale 0-100: ‘Not at all irritated/angry’ to ‘very irritated/angry’<br>[NL] VAS schaal 0-100: ‘Helemaal niet geïrriteerd/boos’ tot ‘heel erg geïrriteerd/boos’ |
| *[EN] Mealtime intensity stressed<br>[NL] Eetmoment intensiteit gestrest | [EN] How stressed did you feel during this meal?<br>[NL] Hoe gestrest voelde u zich tijdens dit eetmoment? | [EN] VAS scale 0-100 0-10; ‘Not at all stressed’ to ‘very stressed’<br>[NL] VAS schaal 0-100 0-10; ‘Helemaal niet gestrest’ tot ‘heel erg gestrest’ |
| *[EN] Mealtime intensity relaxed<br>[NL] Eetmoment intensiteit ontspannen | [EN] How relaxed/calm did you feel during this meal?<br>[NL] Hoe ontspannen/kalm voelde u zich tijdens dit eetmoment? | [EN] VAS scale 0-100: ‘Not at all relaxed/calm’ to ‘very relaxed/calm’<br>[NL] VAS schaal 0-100: ‘Helemaal niet ontspannen/kalm’ tot ‘heel erg ontspannen/kalm’ |
| *[EN] Mealtime intensity happy<br>[NL] Eetmoment intensiteit blij | [EN] How happy/cheerful did you feel during this meal?<br>[NL] Hoe blij/vrolijk voelde u zich tijdens dit eetmoment? | [EN] VAS scale 0-100: ‘Not at all happy/cheerful’ to ‘very happy/cheerful’<br>[NL] VAS schaal 0-100: ‘Helemaal niet blij/vrolijk’ tot ‘heel erg blij/vrolijk’ |
| *[EN] Mealtime intensity sad<br>[NL] Eetmoment intensiteit droevig | [EN] How sad/gloomy did you feel during this meal?<br>[NL] Hoe droevig/somber voelde u zich tijdens dit eetmoment? | [EN] VAS scale 0-100: ‘Not at all sad/gloomy’ to ‘very sad/gloomy’<br>[NL] VAS schaal 0-100: ‘Helemaal niet droevig/somber’ tot ‘heel erg droevig/somber’ |
| *[EN] Mealtime intensity bored?<br>[NL] Eetmoment intensiteit verveeld | [EN] How bored did you feel during this meal?<br>[NL] Hoe verveeld voelde u zich tijdens dit eetmoment? | [EN] VAS scale 0-100: ‘Not at all bored’ to ‘very bored’<br>[NL] VAS schaal 0-100: ‘Helemaal niet verveeld’ tot ‘heel erg verveeld’ |
| *[EN] Mealtime intensity tired<br>[NL] Eetmoment intensiteit vermoeid | [EN] How tired did you feel during this meal?<br>[NL] Hoe vermoeid voelde u zich tijdens dit eetmoment? | [EN] VAS scale 0-100: ‘Not at all tired’ to ‘very tired’<br>[NL] VAS schaal 0-100: ‘Helemaal niet vermoeid’ tot ‘heel erg vermoeid’ |

| | | |
|---|---|---|
| *[EN] Mealtime intensity energetic<br>[NL] Eetmoment intensiteit energiek | [EN] How energetic did you feel during this meal?<br>[NL] Hoe energiek voelde u zich tijdens dit eetmoment? | [EN] VAS scale 0-100: ‘Not at all energetic’ to ‘very energetic’<br>[NL] VAS schaal 0-100: ‘Helemaal niet energiek’ tot ‘heel erg energiek’ |
| *[EN] Mealtime intensity confident<br>[NL] Eetmoment intensiteit zeflverzekerd | [EN] How confident did you feel during this meal?<br>[NL] Hoe zelfverzekerd voelde u zich tijdens dit eetmoment? | [EN] VAS scale 0-100: ‘Not at all confident’ to ‘very confident’<br>[NL] VAS schaal 0-100: ‘Helemaal niet zelfverzekerd’ tot ‘heel erg zelfverzekerd’ |
| *[EN] Mealtime intensity insecure<br>[NL] Eetmoment intensiteit onzeker | [EN] How insecure did you feel during this meal?<br>[NL] Hoe onzeker voelde u zich tijdens dit eetmoment? | [EN] VAS scale 0-100: ‘Not at all insecure’ to ‘very insecure’<br>[NL] VAS schaal 0-100: ‘Helemaal niet onzeker’ tot’ heel erg onzeker’ |
| *[EN] Mealtime intensity ashamed<br>[NL] Eetmoment intensiteit beschaamd | [EN] How embarrassed/guilty did you feel during this meal?<br>[NL] Hoe beschaamd/schuldig voelde u zich tijdens dit eetmoment? | [EN] VAS scale 0-100: ‘Not at all ashamed/guilty’ to ‘very ashamed/guilty’<br>[NL] VAS schaal 0-100: ‘Helemaal niet beschaamd/schuldig’ tot ‘heel erg beschaamd/schuldig’ |
| *[EN] Mealtime intensity lonely<br>[NL] Eetmoment intensiteit eenzaam | [EN] How lonely did you feel during this meal?<br>[NL] Hoe eenzaam voelde u zich tijdens dit eetmoment? | [EN] VAS scale 0-100: ‘Not at all lonely’ to ‘very lonely’<br>[NL] VAS schaal 0-100: ‘Helemaal niet eenzaam’ tot ‘heel erg eenzaam’ |
| *[EN] Mealtime: who<br>[NL] Eetmoment: wie | [EN] Who were you with when you ate this?<br>[NL] Met wie was u toen u dit at? | [EN] No one; Partner; Friends; Family members/housemates; Other family members; Colleagues/classmates; Strangers; Other {open text}<br>[NL] Niemand; Partner; Vrienden; Gezinsleden/huisgenoten; Overige familieleden; Collega’s/klasgenoten; Onbekenden; Anders {open tekst} |

| | | |
|---|---|---|
| *[EN] Mealtime: where<br>[NL] Eetmoment: waar | [EN] Where were you when you ate this?<br>[NL] Waar was u toen u dit at? | [EN] At home; Guest at someone's house; Work/school; Restaurant/café; On the road/on the go; Shop/supermarket; Gym/club; Outdoors; Other {open text}<br>[NL] Thuis; Te gast bij iemand; Werk/school; Restaurant/café; Onderweg; Winkel/supermarkt; Sportschool/vereniging/club; Buiten; Anders {open tekst} |
| *[EN] Mealtime: other activities<br>[NL] Eetmoment: andere bezigheden | [EN] Were you doing anything else while you ate this?<br>[NL] Was u nog iets anders aan het doen terwijl u dit at? | [EN] No; Reading (book, newspaper, news); Scrolling on social media; Watching TV; Playing games; Working/studying; Talking; Cooking; Other {open text}<br>[NL] Nee; Lezen (boek, krant, nieuws); Scrollen op sociale media; TV kijken; Spelletje spelen; Werken/studeren; Praten; Koken; Anders {open tekst} |
| [EN] Drinks<br>[NL] Drinken | [EN] Have you had anything to drink in the past 2 hours?<br>[NL] Heeft u sinds in de afgelopen 2 uur iets gedronken? | [EN] Yes; No<br>[NL] Ja; Nee |

| | | |
|---|---|---|
| [EN] Drinks: categorie<br>[NL] Drinken: categorie | [EN] What did you drink?<br>[NL] Wat heeft u gedronken? | [EN] Water; Coffee/tea without sugar/milk; Coffee/tea with sugar/milk; Sweetened coffees; Diet (soft) drinks; Regular (soft) drinks/energy drinks/sports drinks; Juices/smoothies; Dairy drinks (plant-based); Meal shake/protein shake; Alcoholic beverages; Non-alcoholic alternatives<br>[NL] Water; Koffie/thee zonder suiker/melk; Koffie/thee met suiker/melk; Zoete koffies; Light (fris)dranken; Reguliere (fris)dranken/energiedranken/sportdranken; Sappen/smoothies; Zuiveldrank (plantaardig); Maaltijdshake/proteïneshake; Alcoholische drank; Non-alcoholische alternatieven |

#### Sleep

*this 'sleep survey' triggered once per day in the morning as part of the baseline and post-intervention (duplicates) 3-week EMA measurement period.

| Variable | Item | Answer options |
|---|---|---|
| [EN] Seep duration<br>[NL] Slaapduur | [EN] How long did you sleep last night?<br>[NL] Hoe lang heeft u afgelopen nacht geslapen? | [EN] Less than 2 hours; 2 to 5 hours; 5 to 7 hours; 7 to 9 hours; 9 to 12 hours; More than 12 hours<br>[NL] Minder dan 2 uur; 2 tot 5 uur; 5 tot 7 uur; 7 tot 9 uur; 9 tot 12 uur; Meer dan 12 uur |
| [EN] Sleep quality<br>[NL] Slaapkwaliteit | [EN] How rested do you feel?<br>[NL] Hoe uitgerust voelt u zich? | [EN] VAS scale 0-100: 'Not at all rested' to 'very rested'<br>[NL] VAS schaal 0-100: 'Helemaal niet uitgerust' tot 'heel erg uitgerust' |
| [EN] Nightly eating moment<br>[NL] Eetmoment 's nachts | [EN] Did you eat anything between 22:00 and 06:00 last night?<br>[NL] Heeft u vannacht tussen 22:00 en 06:00 iets gegeten? | [EN] Yes; No<br>[NL] Ja; Nee<br>*the following are asked if yes answered here* |
| [EN] Food category<br>[NL] Eetcategorie | [EN] What did you eat? You can select multiple answers. | [EN] Bread/cereal/pancakes; Dairy/egg; Vegetables/fruit; |

| | | |
|---|---|---|
| | [NL] Wat heeft u gegeten? U kunt meerdere antwoorden selecteren. | Soup; Meal; Snack; Pastry/dessert<br>[NL] Brood/ontbijtgranen/pannenkoeken; Zuivel/ei; Groente/fruit; Soep; Maaltijd; Tussendoortje/snack; Gebak/dessert<br>*the following are repeated for the amount eating moments indicated* |
| *[EN] Subcategory: Bread/cereals/pancakes<br>[NL] Subcategorie: Brood/ontbijtgranen/pannenkoeken | [EN] Bread/cereal/pancakes: what did you eat? You can select multiple answers.<br>[NL] Brood/ontbijtgranen/pannenkoeken: wat heeft u gegeten? U kunt meerdere antwoorden selecteren. | [EN] Crackers/rusks/rice wafers (with toppings); Baguette/sandwich (with toppings); Sweet rolls/pancakes/little Dutch pancakes/French toast; Breakfast porridge/oatmeal/muesli (with/without milk); Cruesli/granola (with/without milk); Cornflakes/sweet breakfast cereals (with/without milk)<br>[NL] Cracker/beschuit/rijstwafel (met beleg); (Stok)brood(je)/boterham (met beleg); Zoet brood(je)/pannenkoeken/poffertjes/wentelteefjes; Ontbijtpap/havermout/muesli (met/zonder melk); Cruesli/granola (met/zonder melk); Cornflakes/zoete ontbijtgranen (met/zonder melk) |
| *[EN] Subcategory: Dairy/egg<br>[NL] Subcategorie: Zuivel/ei | [EN] Dairy/eggs: What did you eat? You can select multiple answers.<br>[NL] Zuivel/ei: wat heeft u gegeten? U kunt meerdere antwoorden selecteren. | [EN] (Plant-based) yogurt/quark; Egg<br>[NL] (Plantaardige) yoghurt/kwark; Ei |
| *[EN] Subcategory: Vegetables/fruit<br>[NL] Subcategorie: Groente/fruit | [EN] Vegetables/fruit: what did you eat? You can select multiple answers.<br>[NL] Groente/fruit: wat heeft u gegeten? U kunt meerdere antwoorden selecteren. | [EN] Fruit/fruit salad; (snack/small) vegetables; Side salad; Dried fruit<br>[NL] Fruit/fruitsalade; (Snoep/snack)groente; Bijgerecht salade; Gedroogde vruchten |

| | | |
|---|---|---|
| *[EN] Subcategory: Soup<br>[NL] Subcategorie: Soep | [EN] Soup: What did you eat? You can select multiple answers.<br>[NL] Soep: wat heeft u gegeten? U kunt meerdere antwoorden selecteren. | [EN] Soup; Filled soup/meal soup<br>[NL] Soep; Gevulde soep/maaltijdsoep |
| *[EN] Subcategory: Meal<br>[NL] Subcategorie: Maaltijd | [EN] Meal: What did you eat? You can select multiple answers.<br>[NL] Maaltijd: wat heeft u gegeten? U kunt meerdere antwoorden selecteren. | [EN] Potato-vegetable-meat/fish/meat substitute; Rice/noodle/pasta dish; Couscous/bulgur/quinoa dish; Filled roll/wrap/taco/pita; Quiche/savory pie/homemade pizza; Sushi/tapas; Fries/fried snack/pizza; Meal salad<br>[NL] Aardappel-groente-vlees/vis/vleesvervanger; Rijst/noedel/pasta-gerecht; Couscous/bulgur/quinoa-gerecht; Gevuld broodje/wrap/taco/pita; Quiche/hartige taart/zelfgemaakte pizza; Sushi/tapas; Friet/gefrituurde snack/ pizza; Maaltijdsalade |
| *[EN] Subcategory: Snack<br>[NL] Subcategorie: Tussendoortje/snack | [EN] Snack: What did you eat? You can select multiple answers.<br>[NL] Tussendoortje/snack: wat heeft u gegeten? U kunt meerdere antwoorden selecteren. | [EN] Cereal bar/protein bar/breakfast biscuit/fruit biscuit; Cookie/waffle; Chips/salted toast or nuts/popcorn; Chocolate (bar)/M&Ms/candy; Unsalted nuts/kernels/seeds; Fried snack; charcuterie<br>[NL] Granenreep/proteïnereep/ontbijtkoek/fruitbiscuit; Koekje/wafel; Chips/gezouten toastjes of noten/popcorn; Chocolade(reep)/M&Ms/snoep; Ongezouten noten/pitten/zaden; Gefrituurde snack; kaasjes/worstjes |
| *[EN] Subcategory: Pastry/dessert<br>[NL] Subcategorie: Gebak/dessert | [EN] Pastry/dessert: What did you eat? You can select multiple answers.<br>[NL] Gebak/dessert: wat heeft u gegeten? U kunt meerdere antwoorden selecteren. | [EN] Cake/pastry/pie/candies/profiteroles; Ice cream/ice cream cake; Custard/pudding/dessert; Water ice/sorbet; Cheese board |

| | | |
|---|---|---|
| | | [NL] Taart/gebak/vlaai/bonbons/soesjes; Roomijs/ijstaart; Vla/pudding/dessert; Waterijs/sorbetijs; Kaasplank |
| *[EN] Mealtime time of day<br>[NL] Eetmoment tijd | [EN] What time approximately did you eat this last night? You can select multiple answers.<br>[NL] Hoe laat heeft u dit ongeveer gegeten vannacht? U kunt meerdere antwoorden selecteren. | [EN] Between 10 p.m. and midnight; between midnight and 2 a.m.; between 2 a.m. and 4 a.m.; between 4 a.m. and 6 a.m.<br>[NL] Tussen 22.00 en 00.00; tussen 00.00 en 02.00; tussen 02.00 en 04.00; tussen 04.00 en 06.00 |
| *[EN] Mealtime plan<br>[NL] Eetmoment plan | [EN] Last night I..<br>[NL] Vannacht heb ik… | [EN] Ate much less than I planned; Ate less than I planned; Ate exactly what I planned; Ate more than I planned; Ate much more than I planned<br>[NL] Veel minder gegeten dan ik van plan was; Minder gegeten dan ik van plan was; Precies gegeten wat ik van plan was; Meer gegeten dan ik van plan was; Veel meer gegeten dan ik van plan was |
| *[EN] Mealtime emotion<br>[NL] Eetmoment emotie | [EN] How did you feel when you ate this? You can select up to two answers.<br>[NL] Hoe voelde u zich toen u dit at? U kunt maximaal 2 antwoorden selecteren. | [EN] Scared/anxious; Irritated/angry; Stressed; Relaxed/calm; Happy/cheerful; Sad/gloomy; Bored; Tired; Energetic; Confident; Insecure; Ashamed/guilty; Lonely<br>[NL] Bang/angstig; Geïrriteerd/boos; Gestresst; Ontspannen/kalm; Blij/vrolijk; Droevig/somber; Verveeld; Vermoeid; Energiek; Zelfverzekerd; Onzeker; Beschaamd/schuldig; Eenzaam |
| *[EN] Mealtime intensity scared<br>[NL] Eetmoment intensiteit bang | [EN] How scared/anxious did you feel during this meal?<br>[NL] Hoe bang/angstig voelde u zich tijdens dit eetmoment? | [EN] VAS scale 0-100: ‘Not at all scared/anxious’ to ‘very afraid/anxious’<br>[NL] VAS schaal 0-100: ‘Helemaal niet bang/angstig’ tot ‘heel erg bang/angstig’ |

| | | |
|---|---|---|
| *[EN] Mealtime intensity irritated<br>[NL] Eetmoment intensiteit geïrriteerd | [EN] How irritated/angry did you feel during this meal?<br>[NL] Hoe geïrriteerd/boos voelde u zich tijdens dit eetmoment? | [EN] VAS scale 0-100: ‘Not at all irritated/angry’ to very ‘irritated/angry’<br>[NL] VAS schaal 0-100: ‘Helemaal niet geïrriteerd/boos’ tot ‘heel erg geïrriteerd/boos’ |
| *[EN] Mealtime intensity stressed<br>[NL] Eetmoment intensiteit gestrest | [EN] How stressed did you feel during this meal?<br>[NL] Hoe gestrest voelde u zich tijdens dit eetmoment? | [EN] VAS scale 0-100 0-100; ‘Not at all stressed’ to ‘very stressed’<br>[NL] VAS schaal 0-100 0-100; ‘Helemaal niet gestrest’ tot ‘heel erg gestrest’ |
| *[EN] Mealtime intensity relaxed<br>[NL] Eetmoment intensiteit ontspannen | [EN] How relaxed/calm did you feel during this meal?<br>[NL] Hoe ontspannen/kalm voelde u zich tijdens dit eetmoment? | [EN] VAS scale 0-100: ‘Not at all relaxed/calm’ to ‘very relaxed/calm’<br>[NL] VAS schaal 0-100: ‘Helemaal niet ontspannen/kalm’ tot ‘heel erg ontspannen/kalm’ |
| *[EN] Mealtime intensity happy<br>[NL] Eetmoment intensiteit blij | [EN] How happy/cheerful did you feel during this meal?<br>[NL] Hoe blij/vrolijk voelde u zich tijdens dit eetmoment? | [EN] VAS scale 0-100: ‘Not at all happy/cheerful’ to ‘very happy/cheerful’<br>[NL] VAS schaal 0-100: ‘Helemaal niet blij/vrolijk’ tot ‘heel erg blij/vrolijk’ |
| *[EN] Mealtime intensity sad<br>[NL] Eetmoment intensiteit droevig | [EN] How sad/gloomy did you feel during this meal?<br>[NL] Hoe droevig/somber voelde u zich tijdens dit eetmoment? | [EN] VAS scale 0-100: ‘Not at all sad/gloomy’ to ‘very sad/gloomy’<br>[NL] VAS schaal 0-100: ‘Helemaal niet droevig/somber’ tot ‘heel erg droevig/somber’ |
| *[EN] Mealtime intensity bored<br>[NL] Eetmoment intensiteit verveeld | [EN] How bored did you feel during this meal?<br>[NL] Hoe verveeld voelde u zich tijdens dit eetmoment? | [EN] VAS scale 0-100: ‘Not at all bored’ to ‘very bored’<br>[NL] VAS schaal 0-100: ‘Helemaal niet verveeld’ tot ‘heel erg verveeld’ |
| *[EN] Mealtime intensity tired<br>[NL] Eetmoment intensiteit vermoeid | [EN] How tired did you feel during this meal?<br>[NL] Hoe vermoeid voelde u zich tijdens dit eetmoment? | [EN] VAS scale 0-100: ‘Not at all tired’ to ‘very tired’<br>[NL] VAS schaal 0-100: ‘Helemaal niet vermoeid’ tot ‘heel erg vermoeid’ |
| *[EN] Mealtime intensity energetic<br>[NL] Eetmoment intensiteit energiek | [EN] How energetic did you feel during this meal?<br>[NL] Hoe energiek voelde u zich tijdens dit eetmoment? | [EN] VAS scale 0-100: ‘Not at all energetic’ to ‘very energetic’<br>[NL] VAS schaal 0-100: ‘Helemaal niet energiek’ tot ‘heel erg energiek’ |
| *[EN] Mealtime intensity confident<br>[NL] Eetmoment intensiteit zeflverzekerd | [EN] How confident did you feel during this meal?<br>[NL] Hoe zelfverzekerd voelde u zich tijdens dit eetmoment? | [EN] VAS scale 0-100: ‘Not at all confident’ to ‘very confident’<br>[NL] VAS schaal 0-100: ‘Helemaal niet |

| | | |
|---|---|---|
| | | zelfverzekerd' tot 'heel erg zelfverzekerd' |
| *[EN] Mealtime intensity insecure<br>[NL] Eetmoment intensiteit onzeker | [EN] How insecure did you feel during this meal?<br>[NL] Hoe onzeker voelde u zich tijdens dit eetmoment? | [EN] VAS scale 0-100: 'Not at all insecure' to 'very insecure'<br>[NL] VAS schaal 0-100: 'Helemaal niet onzeker' tot 'heel erg onzeker' |
| *[EN] Mealtime intensity ashamed<br>[NL] Eetmoment intensiteit beschaamd | [EN] How embarrassed/guilty did you feel during this meal?<br>[NL] Hoe beschaamd/schuldig voelde u zich tijdens dit eetmoment? | [EN] VAS scale 0-100: 'Not at all ashamed/guilty' to 'very ashamed/guilty'<br>[NL] VAS schaal 0-100: 'Helemaal niet beschaamd/schuldig' tot 'heel erg beschaamd/schuldig' |
| *[EN] Mealtime intensity lonely<br>[NL] Eetmoment intensiteit eenzaam | [EN] How lonely did you feel during this meal?<br>[NL] Hoe eenzaam voelde u zich tijdens dit eetmoment? | [EN] VAS scale 0-100: 'Not at all lonely' to 'very lonely'<br>[NL] VAS schaal 0-100: 'Helemaal niet eenzaam' tot 'heel erg eenzaam' |
| *[EN] Mealtime: who<br>[NL] Eetmoment: wie | [EN] Who were you with when you ate this?<br>[NL] Met wie was u toen u dit at? | [EN] No one; Partner; Friends; Family members/housemates; Other family members; Colleagues/classmates; Strangers; Other {open text}<br>[NL] Niemand; Partner; Vrienden; Gezinsleden/huisgenoten; Overige familieleden; Collega's/klasgenoten; Onbekenden; Anders {open tekst} |
| *[EN] Mealtime: where<br>[NL] Eetmoment: waar | [EN] Where were you when you ate this?<br>[NL] Waar was u toen u dit at? | [EN] At home; Guest at someone's house; Work/school; Restaurant/café; On the road/on the go; Shop/supermarket; Gym/(sports)club; Outdoors; Other {open text}<br>[NL] Thuis; Te gast bij iemand; Werk/school; Restaurant/café; Onderweg; Winkel/supermarkt; Sportschool/vereniging/club; Buiten; Anders {open tekst} |
| *[EN] Mealtime: other activities<br>[NL] Eetmoment: andere bezigheden | [EN] Were you doing anything else while you ate this?<br>[NL] Was u nog iets anders aan het doen terwijl u dit at? | [EN] No; Reading (book, newspaper, news); Scrolling on social media; Watching TV; Playing games; |

| | | |
|---|---|---|
| | | Working/studying; Talking; Cooking; Other {open text}<br>[NL] Nee; Lezen (boek, krant, nieuws); Scrollen op sociale media; TV kijken; Spelletje spelen; Werken/studeren; Praten; Koken; Anders {open tekst} |

#### Evaluation

*this 'evaluation survey' triggered after on the last day of the baseline and post-intervention (duplicates) 3-week EMA measurement period.

| Variable | Item | Answer options |
|---|---|---|
| [EN] Burdensome<br>[NL] Belastbaarheid | [EN] How burdensome did you experience completing the questionnaires through this app?<br>[NL] Hoe belastend heeft u het invullen van de vragenlijsten via deze app ervaren? | [EN] VAS scale 0-100: 'not at all demanding to 'very demanding'<br>[NL] VAS schaal 0-100 van 'helemaal niet belastend' tot 'heel erg belastend' |
| [EN] Change (eating and exercise) behaviour<br>[NL] Verandering (eet- en beweeg) gedrag | [EN] To what extent has completing these questionnaires influenced your (eating and exercise) behaviour?<br>[NL] In welke mate heeft het invullen van deze vragenlijsten uw (eet- en beweeg)gedrag beïnvloed? | [EN] VAS scale 0-100: 'not at all affected' to 'very affected'<br>[NL] VAS schaal 0-100: 'helemaal niet beïnvloed tot 'heel erg beïnvloed |
| [EN] Open question change behaviour<br>[NL] Open vraag verandering gedrag | [EN] Briefly describe how completing these questionnaires has influenced your behaviour?<br>[NL] Beschrijf kort hoe het invullen van deze vragenlijsten uw gedrag heeft beïnvloed? | [EN] {open text}<br>[NL] {open tekst} |

### Passive Data

#### Garmin

Using a Garmin Vivosmart 4 or 5, we collect data on:

- Sleep
  - Duration
  - Sleep level qualification
  - Sleep quality (Garmin sleep score) (only Vivosmart 5)
- Dailies
  - Steps
  - Distance
  - Heart rate
- Continuous heart rate per minute
- Stress level (Garmin score)
- Body battery (Garmin score)
- Calories burned (Garmin score)
- Epochs (15 minutes summaries of data)

The Garmin smartwatch is worn for a 3-week period at baseline and post-intervention, simultaneously with use of the Avicenna app.

### Post-intervention measurement

#### *Questions for INFO and ILI*

##### Post-intervention measurement of BMI, WHR and any changes in medication or health

| Variable | Item | Response format |
|---|---|---|
| Picture of weight on weighing scale | [EN] Upload the picture you took of your weight on your scale here.<br>[NL] Upload hier de foto die u heeft gemaakt van uw gewicht op uw weegschaal. | [EN] Upload document/picture.<br>[NL] Bestand/foto uploaden. |
| Current weight | [EN] What is your current weight? (kg) Use a period (.) to indicate decimals, NOT a comma (,).<br>[NL] Wat is uw huidige gewicht? (kg) Gebruik een punt (.) om decimalen aan te geven, GEEN komma (,). | [EN} Weight in kg {numeric}<br>[NL] Gewicht in kg {numeriek} |
| Body height | [EN] What is your height? (cm) *As measured at Maastricht University.<br>[NL] Wat is uw lengte? (cm) *Zoals gemeten op de Universiteit van Maastricht. | [EN} Height in cm {numeric}<br>[NL] Lengte in cm {numeriek} |
| Waist circumference | [EN] Measure your waist circumference using the instructions you received. Enter your waist circumference in centimetres below (maximum one decimal place). Use a period (.) to indicate decimals, NOT a comma (,).<br>[NL] Meet uw taille omtrek met behulp van de instructies die u heeft gekregen. Geef hieronder uw taille omtrek in centimeters (maximaal 1 getal achter de komma). Gebruik een punt (.) om decimalen aan te geven, GEEN komma (,). | [EN] Waist circumference in cm {numeric}<br>[NL] Taille omtrek in cm {numeriek} |
| Hip circumference | [EN] Now measure your hip circumference. Enter your hip circumference in centimetres below (maximum one decimal place). Use a period (.) to indicate decimals, NOT a comma (,).<br>[NL] Meet nu uw heupomtrek. Geef hieronder uw heupomtrek in centimeters (maximaal 1 getal | [EN] Hip circumference in cm {numeric}<br>[NL] Heup omtrek in cm {numeriek} |

| | | |
|---|---|---|
| | achter de komma). Gebruik een punt (.) om decimalen aan te geven, GEEN komma (,). | |
| | [EN] Has anything changed in your medication in the past 6 months?<br>[NL] Is er in de afgelopen 6 maanden iets veranderd in uw medicatie? | [EN] Yes, namely {open text}; No; Not applicable<br><br>If yes, namely: When did you start taking this medication? (dd/mm/yyyy)<br>[NL] Ja, namelijk {open tekst}; Nee; Niet van toepassing<br><br>Als ja, namelijk; Wanneer bent u gestart met deze medicatie? (dd/mm/jjjj) |
| Changes in health over the past 6 mo nths | [EN] Did anything change in your health in the past 6 months?<br>[NL] Is er in de afgelopen 6 maanden iets veranderd in uw gezondheid? | [EN] Yes, namely {open text}; No.<br>[NL] Ja, namelijk {open tekst}; Nee. |
| Pregnancy | [EN] Did you become pregnant in the past 6 months?<br>[NL] Bent u in de afgelopen 6 maanden zwanger geraakt? | [EN] Yes, I am/have been pregnant; No; Not applicable.<br>[NL] Ja, ik ben zwanger (geweest); Nee; Niet van toepassing. |

#### Treatment adherence & satisfaction

| Variable | Item | Response format |
|---|---|---|
| Book chapters read | [EN] Which parts of the book have you read? (multiple options possible)<br>[NL] Welke delen van het boek heeft u gelezen? (meerdere opties mogelijk) | [EN] All Chapters; Chapter 1: What's Your Plan?; Chapter 2: Behaviour Change; Chapter 3: Nutrition; Chapter 4: Exercise & Sports; I haven't read any chapters.<br>[NL] Alle hoofdstukken; Hoofdstuk 1: Wat is jouw plan?; Hoofdstuk 2; Gedragsverandering; Hoofdstuk 3: Voeding; Hoofdstuk 4: Beweging & Sport; Ik heb geen hoofdstukken gelezen. |
| Book tasks completed | [EN] Which tasks from the book did you complete?<br>[NL] Welke opdrachten uit het boek heeft u gemaakt? | [EN] All assignments; Almost all assignments; Some assignments; Almost no assignments; No assignments.<br>[NL] Alle opdrachten; Bijna alle opdrachten; Een aantal opdrachten; Bijna geen opdrachten; Geen opdrachten. |
| Created a progress report | [EN] Did you create a progress report, like on page 17 in the 'Slanker' Workbook'?<br>[NL] Heeft u een overzicht gemaakt van uw voortgang, zoals op pagina 17 in het *Slanker* werkboek? | [EN] Yes; No.<br>[NL] Ja; Nee. |

| | | |
|---|---|---|
| Created a behaviour change plan | [EN] Did you develop a behaviour change plan based on the book, following the steps on page 39 of the 'Slanker' Workbook'?<br>[NL] Heeft u op basis van het boek een gedragsveranderingplan opgesteld, volgens de stappen op pagina 39 in het *Slanker* werkboek? | [EN] Yes; No.<br>[NL] Ja; Nee. |
| Created a nutritional plan | [EN] Did you create a nutritional plan based on your energy needs, which you calculated using the method on page 106 of the 'Slanker' Workbook'?<br>[NL] Heeft u een voedingsplan opgesteld op basis van de energiebehoefte die u heeft uitgerekend met de methode op pagina 106 van het *Slanker* werkboek? | [EN] Yes; No.<br>[NL] Ja; Nee. |
| Created an exercise plan | [EN] Did you make an exercise plan based on Chapter 4 of the 'Slanker' Workbook'?<br>[NL] Heeft u op basis van hoofdstuk 4 een beweegplan opgesteld van het *Slanker* werkboek? | [EN] Yes; No.<br>[NL] Ja; Nee. |
| Comprehensiveness of understanding information in workbook | [EN] I found the information in the 'Slanker' Workbook'...<br>[NL] Ik vond de informatie in het Slanker werkboek... | [EN] Likert scale 0-10; 0 = difficult to understand, 10 = easy to understand<br>[NL] Likert schaal 0-10; 0 = moeilijk te begrijpen, 10 = makkelijk te begrijpen |
| Most informative chapters | [EN] Which chapters of the book did you find most informative?<br>[NL] Welke hoofdstukken uit het boek vond u het meest informatief? | [EN] Sort your answers from most to least informative. Chapter 1: What's Your Plan?; Chapter 2: Behaviour Change; Chapter 3: Nutrition; Chapter 4: Exercise & Sports.<br>[NL] Sleep de antwoorden in volgorde van meest naar minst informatief. Hoofdstuk 1: Wat is jouw plan?; Hoofdstuk 2; Gedragsverandering; Hoofdstuk 3: Voeding; Hoofdstuk 4: Beweging & Sport. |
| Most educational chapter | [EN] From which chapters did you learn the most?<br>[NL] Van welke hoofdstukken heeft u het meest geleerd? | EN] Sort your answers from most to least informative for you. Chapter 1: What's Your Plan?; Chapter 2: Behaviour Change; Chapter 3: Nutrition; Chapter 4: Exercise & Sports.<br>[NL] Sleep de antwoorden in volgorde van meest naar minst informatief. Hoofdstuk 1: Wat is jouw plan?; Hoofdstuk 2; |

| | | |
|---|---|---|
| | | Gedragsverandering; Hoofdstuk 3: Voeding; Hoofdstuk 4: Beweging & Sport. |
| Frequency of book use | [EN] How often did you consult the book in the past 6 months?<br>[NL] Hoe vaak heeft u het boek erbij genomen in de afgelopen 6 maanden? | [EN] Never; Once a month; Several times a month; Once a week; Several times a week; Almost daily; Several times a day.<br>[NL] Nooit; Meermaals per maand; Meerdere keren per dag; Eenmaal per week; Meerdere keren per week; Bijna dagelijks; Meermaals per dag. |
| Application of information | [EN] How have you applied the information from the book to your daily life? For example, are there any new habits you've adopted and will continue with, such as cycling to work more often?<br>[NL] Hoe heeft u de informatie uit het boek toegepast in uw dagelijks leven? Bijvoorbeeld: zijn er nieuwe gewoontes die u heeft opgepakt en blijft voortzetten, zoals vaker met de fiets naar uw werk? | [EN] {open text}<br>[NL] {open tekst} |
| Discussion of book with others | [EN] Did you discuss the book's content with people close to you? (for example: friends, family, …?)<br>[NL] Heeft u de inhoud van het boek besproken met mensen uit uw (nabije) omgeving? (bijvoorbeeld: vrienden, familie, …?) | [EN] Yes; No; If yes, with whom did you discuss the book? {open text}<br>[NL] Ja; Nee; Zo ja, met wie heeft u het boek besproken {open tekst} |
| Book score | [EN] If you had to rate this book on a scale of 0 to 10, what rating would you give it?<br>[NL] Als u het boek een cijfer zou moeten geven tussen 0 en 10, welk cijfer zou u dan geven? | [EN] Slider from 0 to 10<br>[NL] Slider van 0 tot 10 |
| Additional comments | [EN] Do you still have any comments about the book?<br>[NL] Heeft u nog opmerkingen over het boek? | [EN] {open text}<br>[NL] {open tekst} |
| Smartphone apps used [only INFO] | [EN] Have you used smartphone apps to track your lifestyle in the past six months? These could be apps that help you with workout schedules or track your eating habits. For example, the FIT.nl PRO app, myfitnesspal, calorie tracker, etc.<br>[NL] Heeft u in de afgelopen 6 maanden gebruik gemaakt van smartphone apps om uw leefstijl te meten? Dit kunnen apps zijn die u helpen met trainingsschema's of uw | [EN] Yes; No.<br>[NL] Ja; Nee. |

| | | |
|---|---|---|
| | eetgedrag bijhouden. Bijvoorbeeld de FIT.nl<br>PRO app, myfitnesspal, calorieën tracker, etc. | |
| Specific apps [only INFO] | [EN] If yes, for the previous question: Which app(s) did you use?<br>[NL] Zo ja, bij vorige vraag; Welke app(s) heeft u gebruikt? | [EN] {open text}<br>[NL] {open tekst} |
| Frequency of app usage [only INFO] | [EN] If yes, to the previous question: How often did you use this app(s)?<br>[NL] Zo ja, bij vorige vraag; Hoe vaak heeft u deze app(s) gebruikt? | [EN] Daily; Several times a week; Weekly; Monthly; Less than monthly.<br>[NL] Dagelijks; Meermaals per week; Wekelijks; Maandelijks; Minder dan maandelijks. |
| Smartwatch use | [EN] Have you used a smartwatch to track your lifestyle in the past 6 months?<br>[NL] Heeft u in de afgelopen 6 maanden gebruik gemaakt van een smartwatch om uw leefstijl te meten? | [EN] Yes, with the smartwatch I received for this study; Yes, with my own smartwatch; No.<br>[NL] Ja, met de smartwatch die ik voor dit onderzoek heb gekregen; Ja, met mijn eigen smartwatch; Nee. |

#### *Additional for ILI*

##### Working Alliance Inventory – Short revised (client) (WAI-SR)

| | | | |
|---|---|---|---|
| Reference | Hatcher, R. L., & Gillaspy, J. A. (2006). Development and validation of a revised short version of the Working Alliance Inventory. *Psychotherapy Research, 16*, 12–25. | | |
| Dutch translation | Stinckens N, Ulburghs A, Claes L. De Werkalliantie als sleutelelement in het therapiegebeuren. Meting met behulp Van De WAV-12: de Nederlandse vertaling Van De Working Alliance. Inventory Tijdschr Klin Psychol. 2009;39:44–60. | | |
| Total number of items | 12 | Subscales | 3 subscales: goal [4 items: 4-6-8-11], task [4 items: 1-2-10-12], bond [4 items: 3-5-7-9] |
| Description | This questionnaire measures three aspects of therapist-client alliance; agreement on tasks, agreement on goals, and development of an affective bond between patient and therapist.<br>Respondents, clients, answer to statements such as: “As a result of these sessions it is clearer as to how I might be able to change.” or “*name of therapist* and I respect each other.” Respondents answer on a 5-point Likert scale ranging from ‘never’ (1) to ‘always’ (5). | | |
| Response format | 1 Never<br>2 Seldom<br>3 Sometimes<br>4 Often<br>5 Always | | |
| Skip logic | None. | | |

| | |
|---|---|
| Scoring | Sum scores of all items are calculated or can be calculated per subscale. Higher scores indicate better quality of alliance or a stronger working alliance.<br>No reverse scoring. |

#### Treatment adherence & satisfaction

| Variable | Item | Response format |
|---|---|---|
| Use of FIT.nl app | [EN] Did you use the FIT.nl app?<br>[NL] Heeft u gebruik gemaakt van de FIT.nl app? | [EN] Yes; No.<br>[NL] Ja; Nee. |
| Purpose of use FIT.nl app | [EN] If yes, to the previous question; If yes, what did you mainly use the app for?<br>[NL] Zo ja, bij vorige vraag; Zo ja, waarvoor heeft u de app vooral gebruikt? | [EN] {open text}<br>[NL] {open tekst} |
| Use of other smartphone apps | [EN] Besides the FIT.nl PRO app, have you used any other apps to track your lifestyle? For example, to track your eating habits or for training schedules.<br>[NL] Heeft u, naast de FIT.nl PRO app, nog gebruik gemaakt van andere apps om uw leefstijl te meten? Bijvoorbeeld om uw eetgedrag bij te houden of voor trainingsschema's. | [EN] Yes; No.<br>[NL] Ja; Nee. |
| Which other smartphone apps | [EN] If yes, for the previous question: Which app(s) did you use?<br>[NL] Zo ja, bij vorige vraag; Welke app(s) heeft u gebruikt? | [EN] {open text}<br>[NL] {open tekst} |
| Frequency of use other smartphone apps | [EN] If yes, to the previous question: How often did you use this app(s)?<br>[NL] Zo ja, bij vorige vraag; Hoe vaak heeft u deze app(s) gebruikt? | [EN] Daily; Several times a week; Weekly; Monthly; Less than monthly.<br>[NL] Dagelijks; Meermaals per week; Wekelijks; Maandelijks; Minder dan maandelijks. |
| Atmosphere group sessions | [EN] How did you experience the atmosphere during the group sessions?<br>[NL] Hoe heeft u de sfeer tijdens de groepssessies ervaren? | [EN] Likert scale from 0-10; 0 = not pleasant, 10 = very pleasant.<br>[NL] Likert schaal van 0-10; 0 = niet prettig, 10 = heel prettig. |
| Interactiveness group sessions | [EN] How interactive were the group sessions?<br>[NL] Hoe interactief waren de groepssessies? | [EN] Likert scale from 0-10; 0 = not interactive, 10 = very interactive.<br>[NL] Likert schaal van 0-10; 0 = niet interactief, 10 = heel interactief. |
| Comfortableness group sessions | [EN] How comfortable did you feel during the group sessions? For | [EN] Likert scale from 0-10; 0 = not comfortable, 10 = very comfortable. |

| | | |
|---|---|---|
| | example, to share personal stories. [NL] Hoe comfortabel voelde u zich tijdens de groepssessies? Bijvoorbeeld om persoonlijke verhalen te delen. | [NL] Likert schaal van 0-10; 0 = niet comfortabel, 10 = heel comfortabel. |
| Score group sessions | [EN] If you had to rate the group sessions on a scale of 0 to 10, what grade would you give them? [NL] Als u de groepssessies een cijfer zou moeten geven tussen 0 en 10, welk cijfer zou u dan geven? | [EN] Scale from 0 to 10. [NL] Schaal van 0 tot 10. |
| Score individual sessions | [EN] If you had to rate the individual sessions with your coach on a scale of 0 to 10, what grade would you give them? [NL] Als u de individuele sessies met uw coach een cijfer zou moeten geven tussen 0 en 10, welk cijfer zou u dan geven? | [EN] Scale from 0 to 10. [NL] Schaal van 0 tot 10. |
| Additional comments about group/individual sessions | [EN] Do you have any comments about the group sessions and/or the individual sessions with your coach? [NL] Heeft u nog opmerkingen over de groepsessies en/of de individuele sessies met uw coach? | [EN] {open text} [NL] {open tekst} |

#### Working Alliance Inventory – Short revised (therapist) (WAI-SR)

| | | | |
|---|---|---|---|
| Reference | Hatcher, R. L., & Gillaspy, J. A. (2006). Development and validation of a revised short version of the Working Alliance Inventory. *Psychotherapy Research, 16*, 12–25. | | |
| Dutch translation | Translated by Anita van Oers & Anne Roefs, 2023. Universiteit Maastricht. | | |
| Total number of items | 10 | Subscales | 3 subscales: goal [3 items: 3, 6, 8] task [3 items: 1, 4, 10], bond [4 items: 2, 5, 7, 9] |
| Description | This questionnaire measures three aspects of therapist-client alliance; agreement on tasks, agreement on goals, and development of an affective bond between client and therapist. Respondents, therapists, answer to statements such as: “We agree on what is important for *name of client* to work on.” or “We are working towards mutually agreed upon goals.” Respondents answer on a 5-point Likert scale ranging from ‘never’ (1) to ‘always’ (5). | | |
| Response format | 1 Never<br>2 Seldom<br>3 Sometimes<br>4 Often | | |

| | |
|---|---|
| | 5 Always |
| Skip logic | None. |
| Scoring | No reverse scoring. |
